# Population-scale trends and variations in human aortic biomechanics

**DOI:** 10.64898/2026.10.09.26365115

**Authors:** Ankush Aggarwal, Ji-Eun Byun, Claudio Capelli, Mark Danton, Nele Famaey, Andrew McBride, Alison Pouch, Silvia Schievano

## Abstract

The biomechanical properties of the human aorta play a key role in cardiovascular function and disease, yet their population-wide trends and variations remain largely unknown. To fill this crucial gap, we quantified variations in aortic size, stiffness and stresses across 57,434 individuals from the UK Biobank using two-dimensional cine MR imaging and a combined deep-learning, probabilistic and biomechanical modelling framework. Aortic diameter and distensibility (inverse of stiffness) were calculated for the ascending and descending aorta, and their conditional distributions were characterised as a function of age, sex, body size and blood pressure. These observations were integrated with ex-vivo tissue mechanics to infer stress distributions across the aortic wall and the associated uncertainty. Our analysis revealed several new relationships. Larger aortas were found to be stiffer, and greater body size (height and/or weight) was associated with larger aortas with no change in stiffness. Aortic distensibility was negatively associated with systolic, but not diastolic, blood pressure. Apparent sex differences in aortic size were substantially reduced after accounting for body size. Importantly, the maximum stress was found to depend not only on aortic size, as previously thought, but also on distensibility, with lower distensibility associated with higher stresses.

The results provide first-of-their-kind population-scale reference distributions for human aortic stiffness, which will improve the personalisation of computational models and digital twins. The correlation between stress and distensibility strongly supports progressing beyond the current diameter-based criteria (based on the law of Laplace) and for the inclusion of stiffness in the clinical risk assessment of aortic diseases. Overall, this study forms the foundation for personalised quantitative medicine and biomechanically informed risk stratification of aortic aneurysms and dissections.

## 1 Introduction

What are the biomechanical properties of the human aorta? In spite of being a simple question, it is far from straightforward to answer. The question is physiologically and clinically relevant, since biomechanics is known to play an important role in many cardiovascular diseases and their treatment [1–3]. For example, the elastic nature of arteries augments the pumping action of the ventricles, especially during the diastolic phase. Indeed, the loss of vascular elasticity (i.e., an increase of stiffness) is associated with higher cardiac load [4]. Arterial stiffness is thus considered to be an indicator of the cardiovascular age [5–7]. Similarly, the stresses in the tissues are sensed by the smooth muscle cells, which in turn regulate the collagen production and thereby the tissue’s elastic properties [8]. Abnormally high/low stresses are hypothesised to lead to tissue degradation [9, 10] and even catastrophic adverse events, such as aortic dissection and rupture [11, 12]. Importantly, stresses vary across the aortic wall thickness. However, this distribution is ignored when the commonly-used law of Laplace is employed, which only provides the average stress value.

On the treatment side, the effectiveness of a medical device is influenced by the interplay between stiffness of blood vessels and that of the device [13–15]. Mismatch can lead to insufficient sealing between the devices and vessel wall, or inability to expand the device during implantation. Furthermore, the stiffness of the vessels may change after the intervention, both in the short- and long-term. These changes can be the determining factors for the procedure’s success [16–18]. While it is possible to know and control the stiffness of the medical devices, the same is not true for human arteries.

One of the challenges in determining the biomechanical properties of arteries is that these are known to vary, across individuals, over time, with location of the artery, and caused by co-morbidities [19, 20]. Thus, determining the average properties is not sufficient; the associated variations must also be quantified and taken into account when making clinical decisions. There is a large body of literature on the biomechanics of arteries which document this variation from two complementary perspectives, each with important limitations.

First, ex-vivo studies provide detailed characterisation of arterial tissue mechanics and constitutive behaviour [21, 22]. However, these studies are typically limited to tens of samples. Moreover, animal tissue is commonly used, which may differ from humans [23, 24]. If the explanted tissue is taken from humans, it is seldom healthy and most likely to be diseased, which has a non-negligible effect [2, 25]. More fundamentally, it is not trivial to connect ex-vivo properties to in-vivo behaviour [26].

Second, time-resolved clinical imaging can characterise the in-vivo state [27], and some of such in-vivo studies have been scaled to large sample sizes, up to 27000 [28]. However, most large studies have limited biomechanics focus and do not provide the stresses. Even for the stiffness, only the univariate (marginal) trends have been investigated [28], which provide limited information and fail to capture nonlinear second-order effects.

This study addresses this crucial knowledge gap regarding the in-vivo biomechanical properties of the human aorta and how these vary across the population. We achieve this by combining the two perspectives and leveraging the unique large-scale imaging dataset provided by UK Biobank [29]. Using cardiac magnetic resonance (CMR) imaging, the objective is to quantify in-vivo biomechanical properties of the human ascending and descending aorta, specifically stiffness and stresses. Particular emphasis is placed on their variation across the population and their multi-variate and nonlinear associations with individual demographic features. By systematically characterising population-level distributions and demographic trends in arterial biomechanics, this study advances our understanding of the factors underlying population-level variation and provides a foundation for next-generation personalised diagnostic and treatment-planning tools.

## 2 Methods

For aortic stiffness, a commonly used metric of distensibility (sometimes also referred to as compliance) was used, which is the inverse of stiffness (i.e., high distensibility means low stiffness, and vice-versa). The methodology employed included automatic segmentation and statistical analysis based on Gaussian mixture model (GMM) (Fig. 1a). Then distensibility, diameter and BP were used to calculate stresses in the arterial wall, while capturing the associated uncertainty (Fig. 1b). These were then analysed using principal component analysis (PCA) and GMM. Each component of the methodological pipeline is described next.

**Fig. 1.**
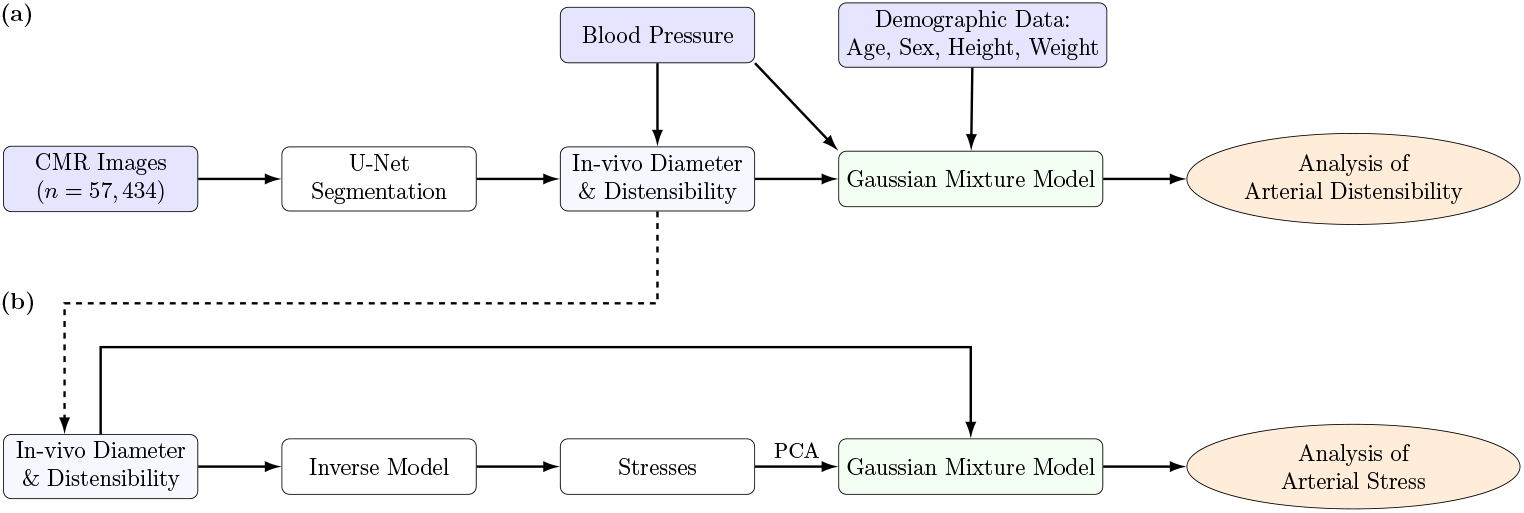
An overview of the methodology for (a) distensibility and (b) stress analyses. Deep-learning segmentation and Gaussian mixture modelling were used for distensibility analysis. Inverse modelling was used for stress calculation capturing the parameter non-uniqueness and the resulting uncertainty in the stresses, which was then analysed using principal component analysis (PCA) and Gaussian mixture modelling.

### 2.1 Image dataset and segmentation

UK Biobank was leveraged, which is a biomedical database from over half a million human volunteers. All data was collected after ethical approval and written informed consent from the participants. A subset of data from participants who underwent CMR was selected (*N* = 66, 970, 48.2% male and 51.8% female, aged 40 to 80 years). The imaging protocol has been described by Petersen et al. [30]. Of particular relevance to this study were the aortic cine images, acquired in the transverse plane at the level of the pulmonary trunk and right pulmonary artery using a balanced steady-state free precession (bSSFP) sequence. In-plane resolution was 1.6mm × 1.6mm, with a slice thickness of 6mm and a temporal resolution of 28 milliseconds.

From each CMR sequence, the ascending and descending aorta was segmented using a previously published U-Net model [28], which was successful for *n* = 57, 434 participants (after quality control checks, i.e., a segmentation is not fragmented and aortic area does not change more than a factor of two). The male/female ratio remained nearly the same (48.8% male and 51.2% female).

### 2.2 Calculation of aortic diameter and distensibility

From the segmented sequence, the cross-sectional luminal areas were calculated, and the maximum and minimum values were considered to be the systolic and diastolic aortic cross-sectional areas (*A*_sys_ and *A*_dias_), respectively. Corresponding diameters (*D*_sys_ and *D*_dias_) were calculated assuming a circular cross-section. In addition, for each subject, their age, sex, height, weight, and blood pressure (BP) measurements were retrieved. The BP values were calculated from the brachial blood pressure by applying a previously described brachial-to-aortic transfer function [31]. Body surface area (BSA) was calculated from height and weight using the Du Bois formula [32]. From the cross-sectional areas and blood pressure measurements, the aortic distensibility δ was derived as

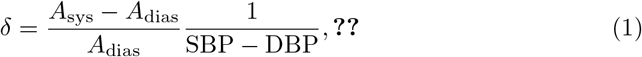

where SBP and DBP are the systolic and diastolic BP values, respectively. All features, their units, and continuous/binary nature are listed in Table 1.

**Table 1.** The characteristics retrieved/derived from UK Biobank (area, diameter and distensibility are each for two locations in the thoracic aorta: ascending and descending)

| Quantity | Units | Nature | Values |
| --- | --- | --- | --- |
| Age | Years | Continuous |  |
| BSA | m<sup>2</sup> | Continuous |  |
| Sex | - | Discrete | Female and male |
| Diastolic pressure (DBP) | mmHg | Continuous |  |
| Systolic pressure (SBP) | mmHg | Continuous |  |
| Diastolic aortic area ($A_{\text{dias}}$) | mm<sup>2</sup> | Continuous | |
| Systolic aortic area ($A_{\text{sys}}$) | mm<sup>2</sup> | Continuous | |
| Diastolic aortic diameter ($D_{\text{dias}}$) | mm | Continuous | |
| Systolic aortic diameter ($D_{\text{sys}}$) | mm | Continuous | |
| Distensibility ($\delta$) | 10<sup>-3</sup>mmHg<sup>-1</sup> | Continuous | |
| Location | - | Discrete | Ascending (Asc) and descending (Dsc) |

### 2.3 Calculation of dependencies and variations

Information theory-based mutual information (MI) was used to quantify dependencies between various characteristics because, unlike linear correlation measures, it captures arbitrary nonlinear dependencies (see Appendix for details on these measures) [33, 34]. The MI was calculated between every pair of continuous variables. MI values with *p <* 0.001 were considered statistically significant (while non-significant MI values were not reported).

To analyse the variations of biomechanical properties (specifically, the cross-sectional area and distensibility), a GMM was fitted to the continuous variables. The number of components and the need for a log-transformation (since all of the variables are constrained to be positive) was determined based on the goodness of the model fit, quantified in terms of the Bayesian information criterion (BIC). The generalisation of the fitted model was confirmed using BIC on a 75/25 train/test split of the dataset. A different GMM was fitted for each subset of discrete variable values (such as, for males/females, and for ascending/descending). From the fitted GMMs, trends were determined by calculating conditional distributions with respect to demographic variables (such as the age, BSA, DBP, and SBP), and plotting their mean and standard deviation values.

The MI does not separate out the second-order effects. For example, the MI between aortic diameter and sex might be non-zero, only because of the difference in the BSA between the two groups (there is a statistically significant difference in male/female BSAs). To remove such second-order effects, conditional mutual information (CMI) was calculated, which describes the mutual information between two quantities, conditioned on a third quantity to be known. In the above example, it will be the CMI between aortic area and sex, conditioned on BSA being known. However, estimating CMI between mixed (continuous and discrete) variables directly from the data is challenging and prone to bias. Therefore, CMI was calculated based on the fitted GMMs, as described in the Appendix.

### 2.4 Calculation of arterial wall stresses

Further to the observable features listed in Table 1 that can be directly analysed, the biomechanical stress in the aortic wall was inversely calculated based on a mathematical model. An innovative approach was used that accounts for associated uncertainties (Fig. 1b), as described next.

#### Biomechanical model

A biomechanical model, described in Appendix, was used to numerically calculate the pressure difference across the arterial wall and the stresses in the wall at a deformed lumen diameter. This computation requires knowledge of the model parameters, which in the model employed herein include nine geometric and material parameters (Table 2). One of the model parameters that affects the resulting stresses is the arterial wall thickness, which cannot be visualised in a CMR. Our strategy to address the lack of knowledge about these model parameters (including wall thickness) is explained next.

**Table 2.** Biomechanical model parameters for ascending and descending aortas and their ranges (taken as mean *±* 2×s.d. from Peña et al. [22]). To capture the full uncertainty, a range of four standard deviations was used.

| Symbol [units] | Description | Ascending | Descending |
| --- | --- | --- | --- |
| $R_i$ [mm] | Internal stress-free radius | [8.65, 13.85] | [8.33, 11.13] |
| $H$ [mm] | Stress-free thickness | [1.6, 3.32] | [1.17, 1.97] |
| $\omega$ [radians] | Opening angle | [0.6, 1.47] | [0.78, 1.82] |
| $\lambda_Z$ [-] | Axial stretch | [1.15, 1.27] | [1.33, 1.61] |
| $\phi$ [°] | Fibre angle | [2.1, 90] | [50.1, 90] |
| $k_2$ [-] | Stress nonlinearity | [0, 8.552] | [0, 27.23] |
| $\kappa$ [-] | Fibre dispersion | [0, 0.33] | [0, 0.33] |
| $\mu$ [kPa] | Isotropic matrix stiffness | n/a | n/a |
| $k_1$ [kPa] | Fibrous stiffness | n/a | n/a |

#### Inverse determination of model parameters

The model parameters were determined inversely based on the known diastolic and systolic areas, *A*_dias_ and *A*_sys_, and blood pressures, SBP and DBP. Importantly, it was recognised that, since the number of unknown parameters is higher than the observed in-vivo data (diastolic and systolic states), the model parameters cannot be uniquely determined. Instead, these were treated as being uncertain, and the resulting uncertainty was propagated to the computed stresses.

While the inverse estimation could be performed using approaches such as Markov Chain Monte Carlo (MCMC), the results for nonlinear models become sensitive to the prior distributions. To resolve this issue, the linear dependence between the luminal blood pressure and stiffness-like parameters (*k*_1_ and *µ*) was exploited. This linearity exists in most biomechanical models, including the one adopted here. Thus, seven parameters (*R*_*i*_, *H, ω, λ*_*z*_, *ϕ, k*_2_ and *κ*), defined in Table 2, were sampled independently from uniform distributions over physiologically plausible ranges based on the ex-vivo study by Peña et al. [22]. Uniform distributions were chosen to minimise prior assumptions regarding the parameter values, while parameter correlations were neglected due to the lack of available population-level data. Consequently, the predicted stresses should be interpreted as distributions conditioned on the assumed physiologically plausible parameter space, rather than unique patient-specific estimates.

For each sampled parameter set, denoted as ***θ***_s_, the remaining two parameters (*µ* and *k*_1_) were determined by enforcing consistency with the observed in-vivo diastolic and systolic states. Specifically, the internal pressures at diastolic and systolic arterial sizes were calculated at two hypothetical parameter sets (*µ* = 1, *k*_1_ = 0) and (*µ* = 0, *k*_1_ = 1). That is, the following four pressure values were evaluated:

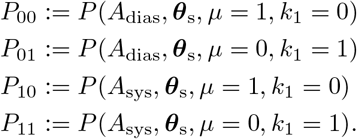

Based on these pressure values, the values of *µ* and *k*_1_ were chosen such that the model-predicted internal pressures at the diastolic and systolic radii matched the recorded diastolic and systolic blood pressures, respectively. That is, the following linear system of equations was solved

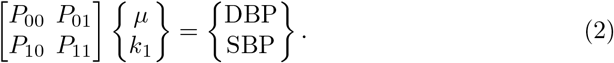

Parameter sets that resulted in a negative stiffness (either *µ <* 0 or *k*_1_ *<* 0) were discarded. Once all model parameters were determined, Cauchy stresses in the three normal directions (radial, circumferential, and axial) at both the diastolic and the systolic configurations were computed at ten equispaced points across the arterial wall thickness. The stress values are expected to have a strong correlation with the BPs, which causes ill-conditioning in the further analysis steps. To address this issue, the stresses were normalised with respect to the lumen BP (diastolic or systolic, as appropriate).

#### Statistical description of stresses

The stress distribution across the arterial wall entails a high 60-dimensional vector. To make the analysis feasible, its dimension was reduced using PCA. Next, a GMM was fitted to the generated samples of in-vivo states and the arterial wall stresses. Spearman’s rank correlations and MI were calculated between the maximum stress values and in-vivo observables. The fitted GMM was then conditioned with respect to varying in-vivo states to analyse the trends, quantified in terms of the mean and standard deviation in the wall stresses.

## 3 Results

### 3.1 Aortic distensibility

From the segmented images, aortic size (quantified in terms of diastolic diameter) and aortic stiffness, (quantified in terms of distensibility, inverse of stiffness) were successfully derived for the ascending and descending aortas in *n* = 57, 434 human volunteers.

#### 3.1.1 Dependencies between observed quantities

From the observed quantities, the distribution of every feature against another is visualised in Fig. 2. There is a co-dependence between almost all variables to varying degrees. Moreover, many of the univariate distributions, such as distensibilities, resemble log-normal. The distributions are split by sex, which show sex-based differences, particularly between the BSA and aortic diameters. Some of the correlations also look nonlinear, indicating the need for MI to quantify their co-dependence.

**Fig. 2.**
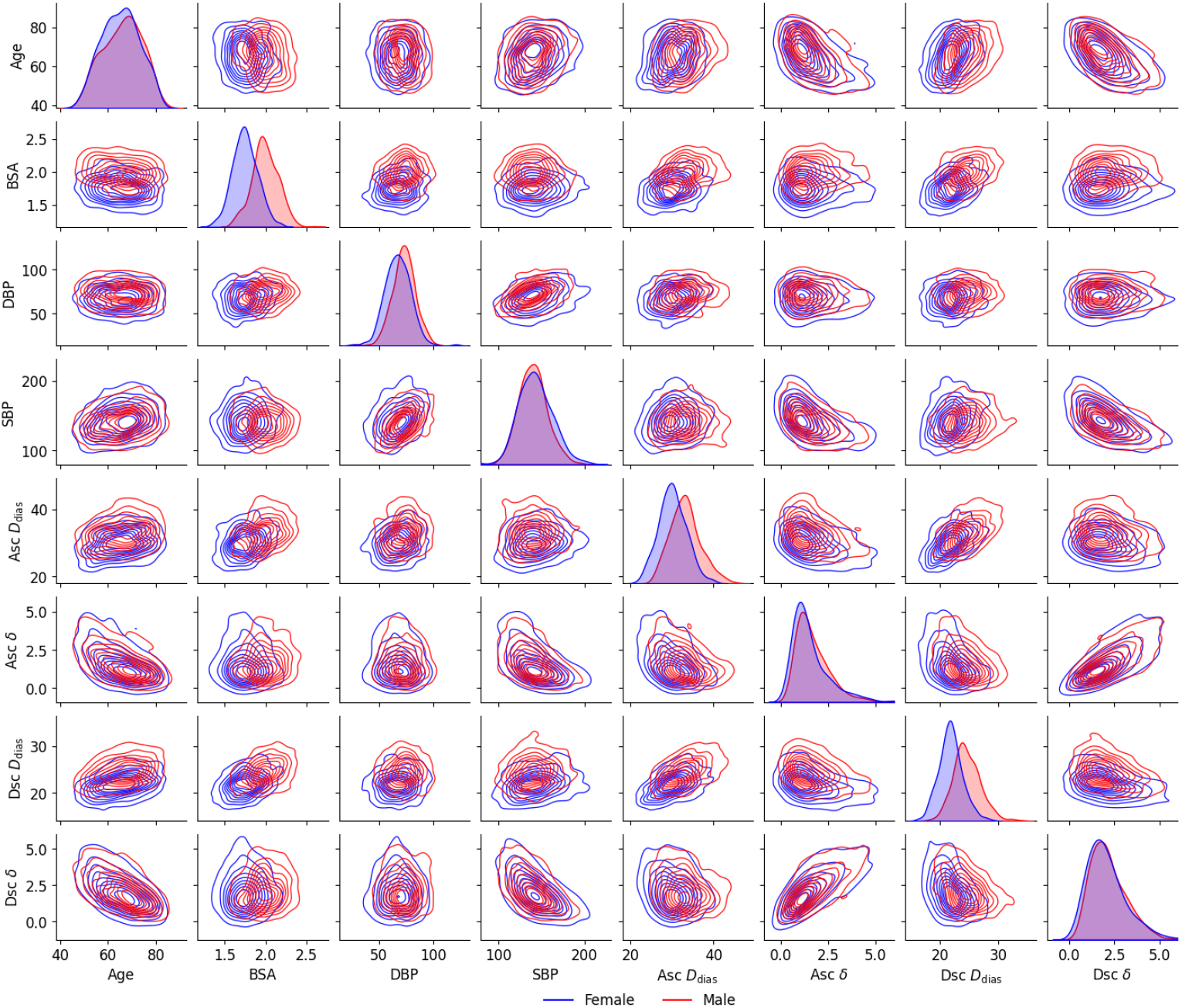
Pair-plot of the continuous features divided into males and females. All variables show some level of correlation, which is quantified and analysed further. There are sex-based differences in the BSA and aortic diameters.

Thus, the MI between all continuous features is shown in Fig. 3. Since all features have statistically significant MI with at least one other feature, none of the features can be considered as independent. There is a strong dependence between the ascending and descending aortic properties (MI of 0.39 and 0.61). Similarly, there is a moderate dependence between aortic size and distensibility, for both ascending and descending aorta (MI of 0.12 and 0.13). The distensibilities depend on SBP (MI of 0.32 and 0.40), but not on DBP (insigificant MI). The aortic size is minimally dependent on the BPs (MI ranging from 0.03 to 0.06). The aortic sizes and distensibilities also depend on age and BSA, although to varying degrees. However, due to the multi-dimensionality of the feature space, the two-dimensional contours and MI provide limited information.

**Fig. 3.**
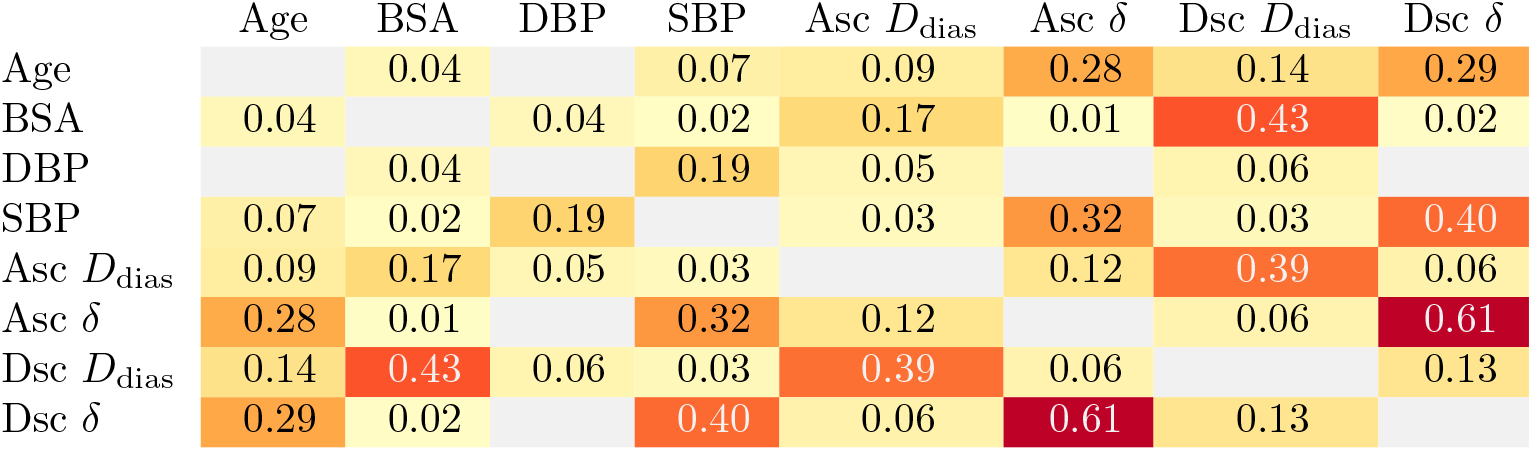
Dependence between continuous features quantified with mutual information (in units of bits), which have been colour-coded to indicate the high dependence variables. Only statistically significant (*p <* 0.001) are shown. Aortic distensibility (δ) depends on all features except the DBP, and there is a high correlation between ascending (Asc) and descending (Dsc) aortic diameters and distensibilities.

#### 3.1.2 Gaussian mixture model fitting and trend inference

A GMM with three components was able to fit multi-dimensional data of six variables (age, BSA, DBP, SBP, aortic size, and aortic distensibility), with a log-transform providing a significant improvement in the fit quality. This applied to the data from ascending/descending and males/females resulted in four fitted GMMs.

We start by analysing the relation between aortic size and distensibility (which showed moderate MI values). For the mean age, BSA and BPs, we plot the variation of aortic distensibility versus diameter (Fig. 4). The negative correlation indicates that larger aortas have lower distensibility (i.e., higher stiffness). This is true for both ascending and descending and for males and females. However, the trend is nonlinear, with the rate of decrease in distensibility slowing down at larger diameters.

**Fig. 4.**
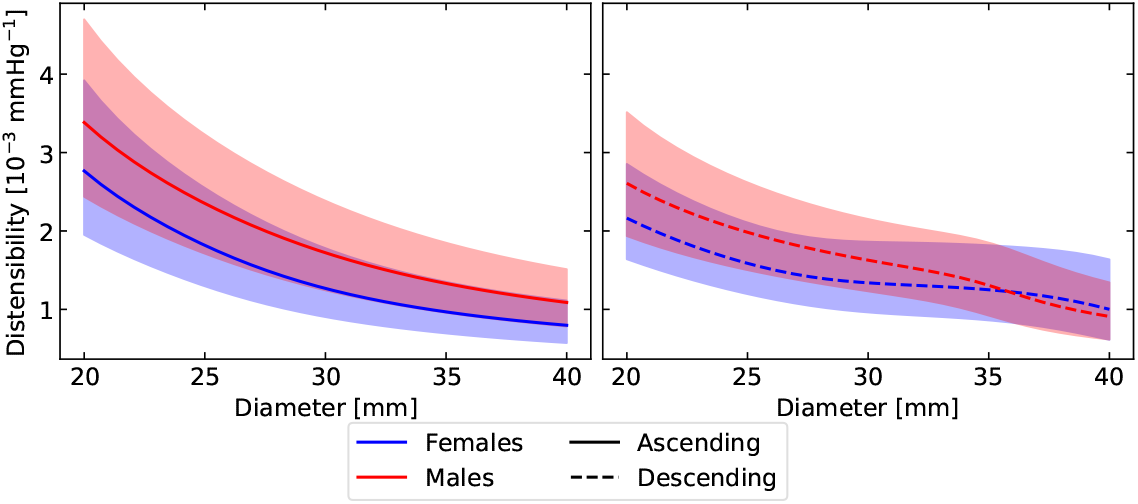
Variation of aortic distensibility as its diameter is varied (with age, body surface area and blood pressures fixed at their respective mean values). Solid/dashed lines indicate mean values and shaded area mark one standard deviation. A consistent trend was found that larger aortas are associated with lower distensibility (stiffer) – in both male/female and ascending/descending aortas.

Given the co-dependence between the aortic size and stiffness, we plot the joint distribution of aortic diameter and distensibility, conditioned on different combinations of age, BSA, and BPs (Fig. 5 and see animated plots in the supplementary material). There is an obvious variation in both diameter and distensibility. While there is a clear difference between ascending and descending aorta diameters, the difference in their distensibilities is less evident. The univariate histograms of aortic sizes showed difference between males and females (Fig. 2). However, the difference in contour distributions for males and females for the same age/BSA/pressures is less obvious. This was confirmed by calculating the CMI between aortic size and sex given the age and BSA, which was 0.009 for ascending and 0.035 for the descending. Similarly, the CMI between aortic distensibility and sex given age and BSA were 0.012 for ascending and 0.007 for descending.

**Fig. 5.**
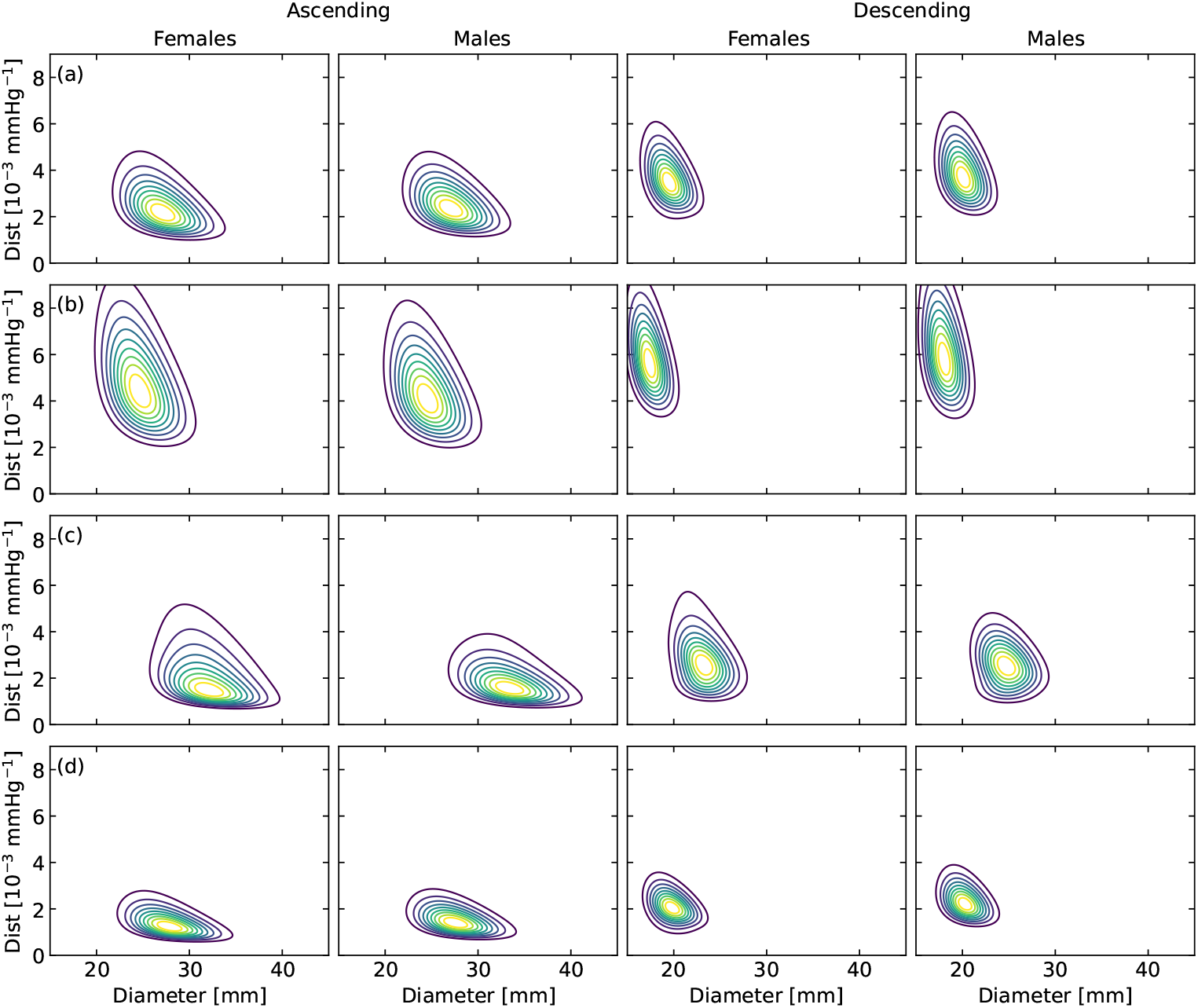
Example conditional distributions of aortic size and distensibility for different feature value combinations: a) age = 65 years, BSA = 1.2 m^2^, DBP = 80 mmHg, SBP = 120 mmHg; b) age = 45 years, BSA = 1.2 m^2^, DBP = 80 mmHg, SBP = 120 mmHg; c) age = 75 years, BSA = 1.8 m^2^, DBP = 80 mmHg, SBP = 120 mmHg; d) age = 65 years, BSA = 1.2 m^2^, DBP = 80 mmHg, SBP = 150 mmHg. The contour lines represent increments in 10 percentile (from 10 to 90). Animated contour plots for varying features are provided as supplementary information.

The trends of aortic size and stiffness is further investigated by systematically varying one of the demographic parameters: age, BSA, DBP, and SBP (Fig. 6). The results highlight important trends. The ascending and descending diameters are clearly different, but their distensibilities are much similar.

**Fig. 6.**
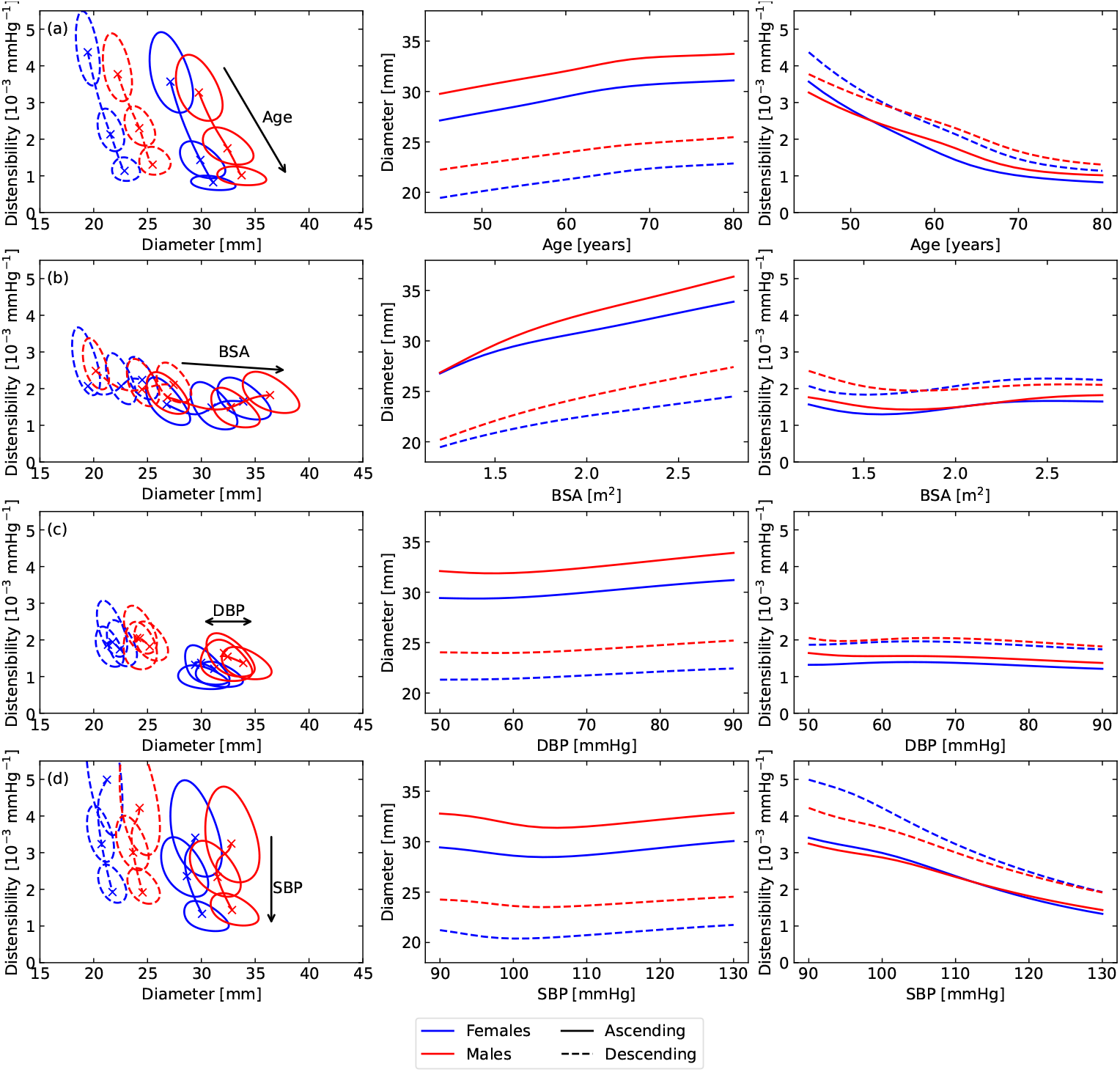
Variation of aortic size and distensibility versus (a) age, (b) BSA, (c) DBP, and (d) SBP. Older humans have larger and stiffer aorta. For humans with larger BSA, the aorta is larger without a change in distensibility. DBP has minimal effect on the aortic properties. Those with higher SBP have a stiffer aorta, without change in the size. Lines represent the mean values, and contour in the left column indicates top 20 percentile range (to highlight the associated variation).

There is a clear increase in the aortic size with age, while the aortic distensibility decreases. The rates of these changes is largely same for both males/females and ascending/descending aortas. Moreover, the trends are nonlinear, especially for distensibility.

With respect to BSA, only the aortic size increases, while the distensibility remains largely unchanged. The rates of change with respect to BSA are slightly different between males and females, with female aortic sizes increasing at a lower rate than males’.

The variation with respect to DBP is almost zero. In contrast, the SBP is associated with a clear change in aortic stiffness (but not the aortic diameter). Specifically, larger SBP is associated with stiffer aortas, and the rate of that change is roughly the same for both ascending/descending and males/females. The only difference seems to be for descending aortas in females at low SBPs.

### 3.2 Aortic stresses

Stresses in the aortic wall were successfully calculated for all samples. Each sample included the normalised (dimensionless) stress distributions at both diastole and systole in three directions (radial, circumferential and longitudinal). This resulted in a combined stress vector of length 60 for each sample. In addition to stresses, the inversely calculated wall thickness was also recorded for further analysis.

The mean and standard deviation of inversely calculated stresses across all samples is shown in Fig. 7. The distributions are plotted against the position across the arterial wall, with 0 depicting the inner (luminal) surface and 1 depicting the outer surface. The radial stress is compressive with value ranging from -1 at the lumen surface to 0 at the exterior surface. The circumferential stress is largely tensile, with normalised value reaching as high as 100+ at the inner surface in some cases. The longitudinal stress varies from compressive to tensile region (up to normalised values of approximately 40).

**Fig. 7.**
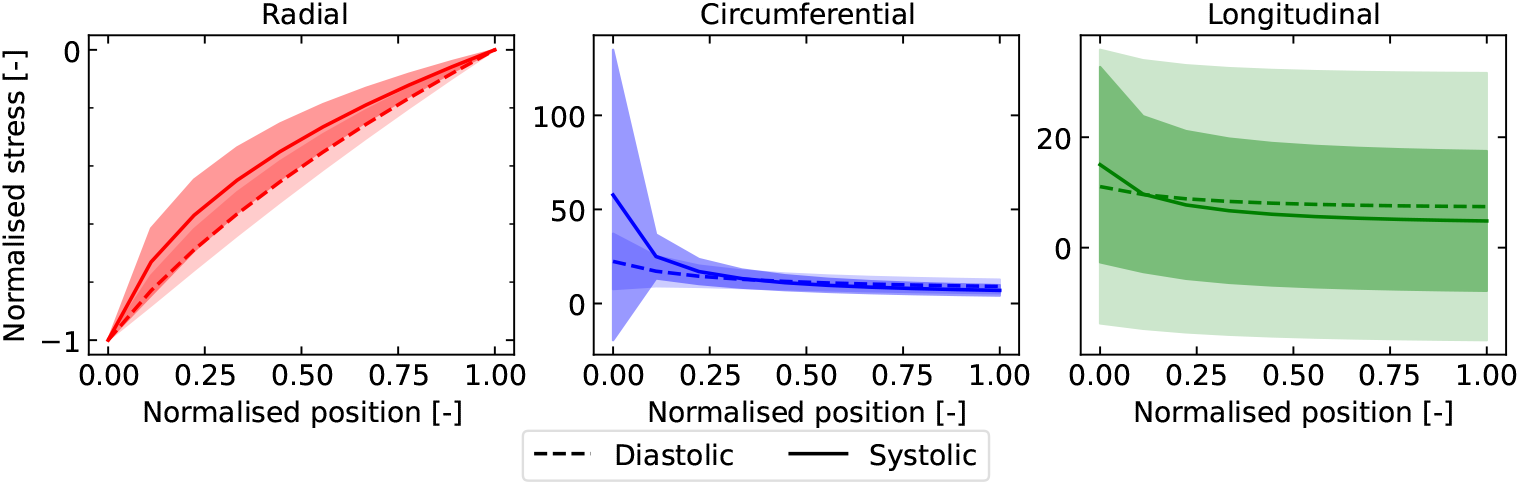
Distribution of the normal, circumferential, and longitudinal stresses (normalised to the blood pressure) in the ascending aorta across the arterial wall (normalised position, 0 being the inner surface and 1 being the outer), with variation for all cases combined (i.e., marginalised to the aortic parameters). Solid lines represent the mean values and shaded areas indicate one standard deviation. Radial stresses are compressive with small uncertainty, while the circumferential and longitudinal stresses are predominantly tensile with high uncertainties.

#### 3.2.1 Relation of stresses and aortic properties

To further analyse how the stresses depend on the aortic properties (such as size, distensibility and BPs), PCA helped reduce the dimension of the stress vectors to six while preserving an explained variance of 0.98+ (two modes for the diastolic stress and four modes for the systolic stress). A GMM with six components was able to fit the ten-dimensional data (aortic size, distensibility, DBP, SBP, and six PCA modes for the stresses). Since the PCA projections can be both positive and negative, log-transform was not applied to those.

The stress distributions (mean values and the quantified uncertainty) conditioned on different values of aortic diameter, distensibility, and BPs are plotted (Fig. 8 and see animated plots in the supplementary material). The stress distributions qualitatively remain the same, but there are quantitative variations. The variation in radial stress with respect to the patient features is almost negligible. In contrast, the circumferential and longitudinal stresses and the associated uncertainty are variable, with both the mean values and uncertainty changing with different inputs.

**Fig. 8.**
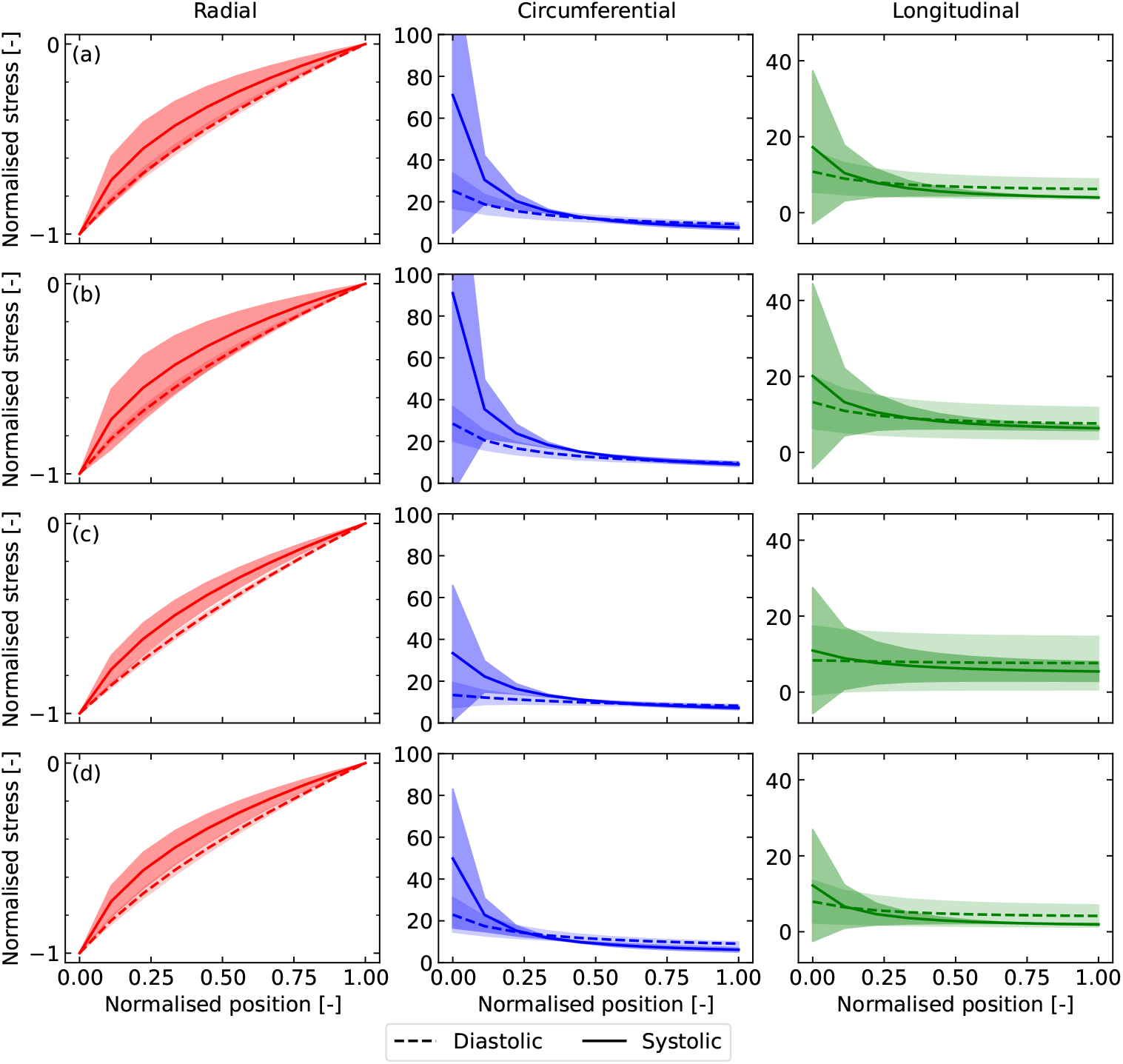
Example conditional distributions of the stresses (normalised to the blood pressure) across the descending arterial wall (normalised position, 0 being the inner surface and 1 being the outer) for different blood pressures, aortic size and distensibility: a) DBP = 65 mmHg, SBP = 130 mmHg, diameter = 35 mm, dist = 2 × 10^−3^ mmHg^−1^; b) DBP = 80 mmHg, SBP = 130 mmHg, diameter = 40 mm, dist = 3 × 10^−3^ mmHg^−1^; c) DBP = 55 mmHg, SBP = 120 mmHg, diameter = 30 mm, dist = 4 × 10^−3^ mmHg^−1^; d) DBP = 75 mmHg, SBP = 150 mmHg, diameter = 30 mm, dist = 1 × 10^−3^ mmHg^−1^. Solid/dashed lines are the mean values and shaded areas represent one standard deviation. Animated plots for varying inputs are provided as supplementary information.

The radial stress distributions are constrained by the boundary conditions; the radial stress must be equal to the lumen BP at the inner surface, and go to zero at the outer surface. Thus, there is no uncertainty in the radial stress at those two positions. However, there is a small uncertainty in the interior of the arterial wall.

The circumferential stresses show a large gradient; the inner surface always carries the highest circumferential stress, which decreases across the wall. Unlike the radial stress, the circumferential stress is non-zero at the external surface. The circumferential stresses also carry a large uncertainty, which is highest at the inner surface but decreases across the wall. Comparing the diastolic and systolic states, the circumferential stress is higher at systole, even after being normalised by the blood pressure.

The highest uncertainty is evident for the longitudinal stresses, which in some cases can have a standard deviation of twice as much as the mean value. This is related to the fact that the in-vivo data used here is from a cross-sectional plane, thus providing little information about the out-of-plane longitudinal deformation or stresses. The mean longitudinal stress values are of the same order of magnitude as the circumferential stress (although generally lower than the circumferential stress at the inner wall). However, the large uncertainty implies that the longitudinal stress may be compressive in some cases. Also, the variation of longitudinal stresses across the arterial wall is relatively small, with close to uniform stress distribution.

From the conditioned distributions, the variation of maximum circumferential and longitudinal stresses versus DBP, SBP, aortic diameter and distensibility are plotted (Fig. 9). The variations in normalised stresses versus DBP and SBP are minimal. However, we find that the maximum stresses increase with diameter, but decrease with distensibility. The results for descending aorta are qualitatively similar, and therefore deferred to the Appendix.

**Fig. 9.**
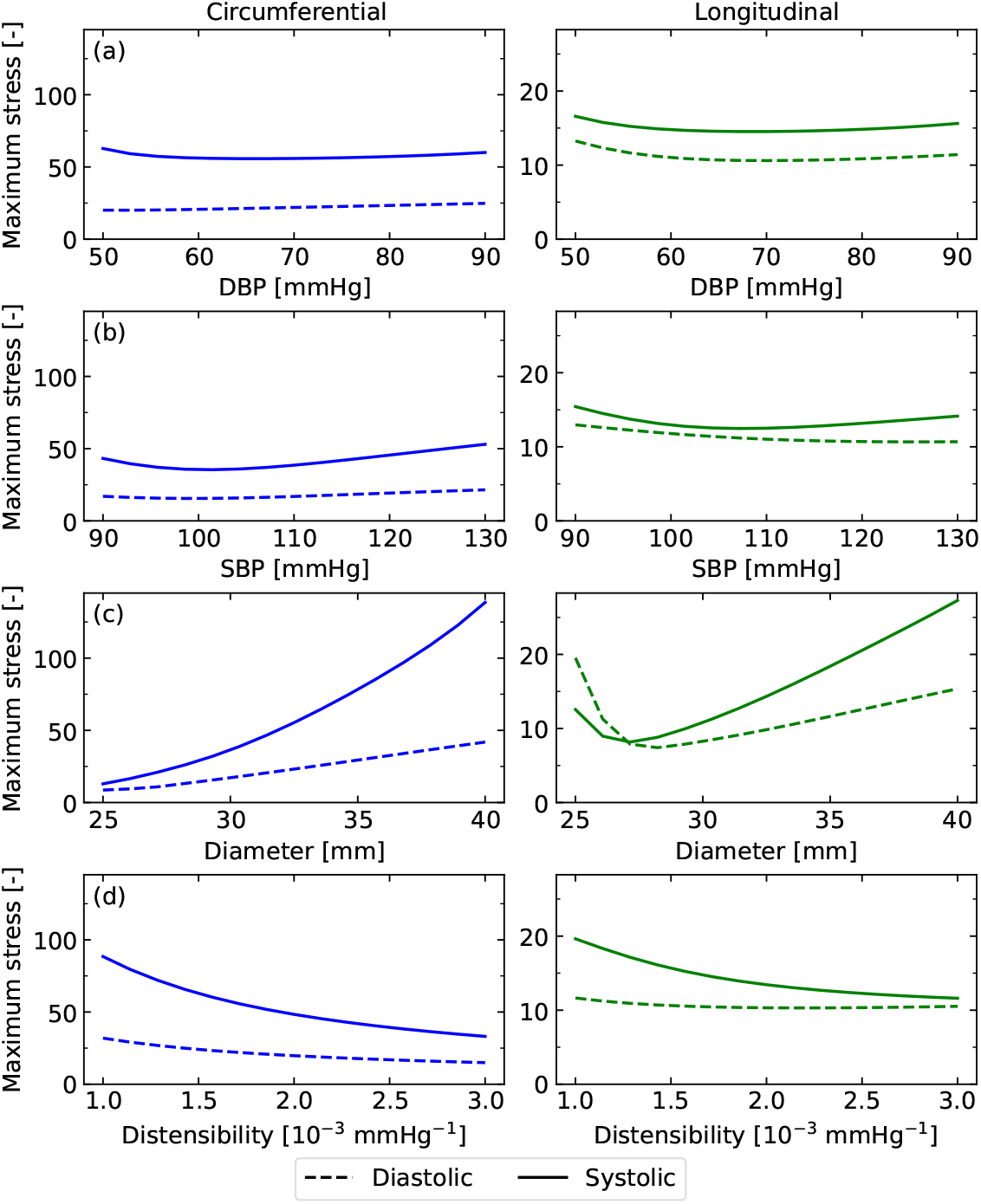
Variation of the expected maximum stress (normalised) as the pressures, size and distensibility of the ascending aorta are varied. Pressures have a minimal effect, however the stress increases with diameter, but decreases with distensibility.

#### 3.2.2 Dependence between observable quantities and stresses

To analyse the dependence between stresses and observable quantities, we look at the mean and maximum stress values, and how these correlate with the aortic diameter, distensibility and wall thickness. Law of Laplace is commonly used to estimate the circumferential stress, which assumes the stress to be uniform across the arterial wall. That is, the mean circumferential stress calculated here is equivalent to the stress given by the law of Laplace, and it was found to be positively correlated with the aortic size, and negatively correlated with the aortic distensibility and wall thickness (Fig. 10).

**Fig. 10.**
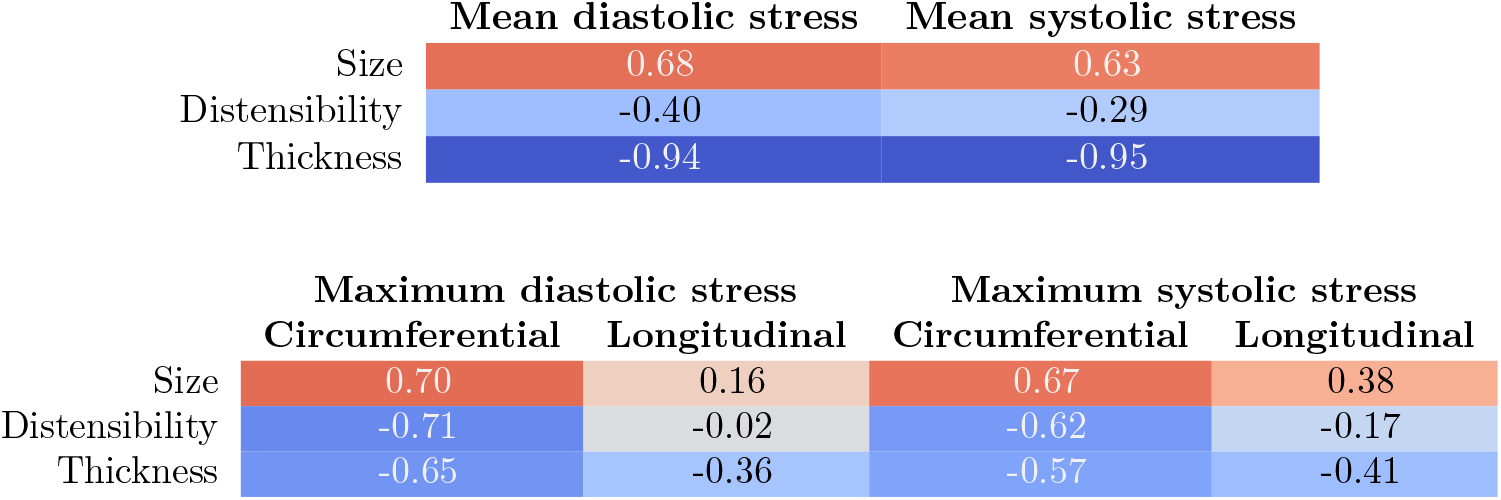
Spearman correlation coefficients between the aortic parameters and mean (top) and maximum (bottom) stresses. Values are colour-coded to highlight negative (blue) and positive (orange/red) correlations. Size is positively correlated with the stresses, while distensibility and thickness are negatively correlated. The correlations with longitudinal stress are small, however.

Biomechanically, the maximum stress values are more relevant than the mean values, as these can lead to wall deterioration, dissection or rupture. Thus, the MI and correlation coefficients between maximum normalised stress values and in-vivo features were calculated (Fig. 10 and 11). Compared to the mean stress, the correlations of maximum circumferential stresses are higher (almost double) with distensibility, but lower with thickness. The MI between the aortic size and maximum circumferential stresses is high, and they have a positive correlation. In contrast, the MI and correlation between the aortic size and maximum longitudinal stresses is relatively lower, although slightly higher at systole than diastole. The correlation between distensibility and maximum circumferential stresses is negative, indicating that more distensible (i.e., compliant) arteries experience lower stresses. The co-dependence between distensibility and circumferential stresses was further confirmed by calculating the CMI between distensibility and maximum circumferential stress, for given aortic size and thickness, which was found to be 0.24 and 0.14 for the diastolic and systolic states, respectively.

**Fig. 11.**
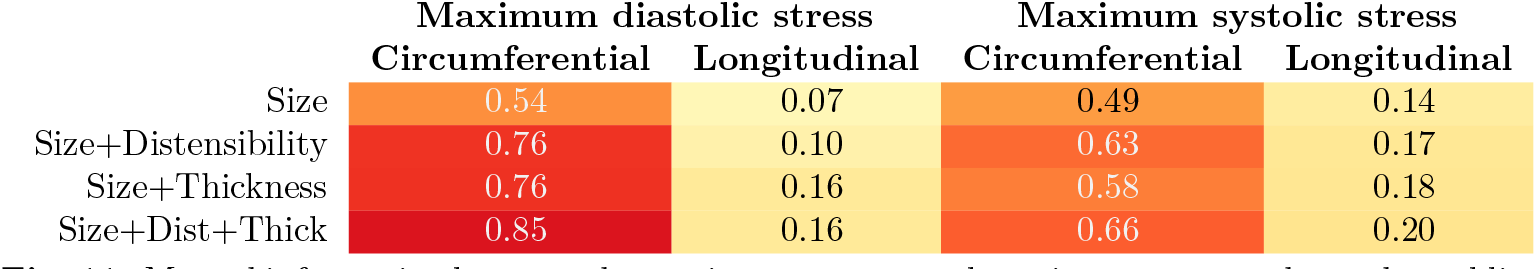
Mutual information between the aortic parameters and maximum stresses shows that adding distensibility and/or thickness to the size can provide us higher information and therefore confidence about the arterial stresses.

The arterial wall thickness, although not observable in the CMR, was determined inversely, and it was found to have a negative correlation with both circumferential and longitudinal stresses. Currently, in the clinical practice, only aortic size/diameter is used to assess the risk of adverse events, which originate from biomechanical stresses. Indeed, this is supported by the high MI between aortic size and maximum circumferential stresses. However, if one would also use aortic distensibility and/or aortic wall thickness values, the MI with circumferential stresses increases by 20–40%. It was found that only the correlations/MI with circumferential stress are high, while those with the longitudinal stress are generally smaller (except with the wall thickness).

## 4 Discussion

There is a large body of evidence on how important a role biomechanics plays in aortic health and diseases [1, 35, 36]. Yet, determining the in-vivo biomechanical properties in the human aorta remains a challenge. This is even more true for healthy population, as most of the clinical data comes from patients [27, 37, 38]. Moreover, the ex-vivo and in-vivo properties differ substantially, and care must be taken when relating or translating them [39–41]. Previous studies have used animal models as surrogates or been limited to small sample sizes (usually tens to hundreds of sample size), which limits the analysis and therefore the relevant insight. The present study fills this important gap by leveraging the unique UK Biobank, with in-vivo data from almost 60 thousand humans from the wider population. The analysis performed led to many crucial and novel insights, which are discussed next.

### 4.1 Main findings on aortic stiffness

It was observed that the aortic stiffness varies substantially across humans, and this variation was quantified. At the same time, this variation in aortic stiffness follows consistent trends. The analysis showed that larger aortas are, on average, stiffer (Fig. 4). Both aortic size and stiffness were found to increase with age (i.e., distensibility decreases with age) (Fig. 6a). This is generally accepted as part of the ageing process [42, 43], and partly responsible for increasing risk of cardiovascular diseases. In contrast, with larger height/weight (combined into BSA), the aorta size increases, but its stiffness does not change appreciably (Fig. 6a). In many clinical guidelines, the aortic size is indeed indexed with respect to the BSA [44–46]. The effect of blood pressure showed interesting trends, whereby the DBP had no correlation with aortic size or stiffness (Fig. 6c). However, with higher SBP, the aortic stiffness increased while the aortic size remained unchanged (Fig. 6d). Though it is challenging to establish whether higher SBP leads to larger stiffness or larger stiffness leads to higher SBP. We note that increasing SBP for a fixed DBP and *A*_dias_ is expected to increase the systolic lumen area *A*_sys_. That is, by definition (Eq. **??**), it is not obvious how δ will vary with SBP. The fact that the MI between SBP and aortic size (both at diastole and systole) was close to zero, there may be regulatory mechanisms that are responsible for adapting the stiffness such that the aortic size remains invariant with respect to any BP variations.

Both ascending and descending and male and female aortas followed these trends. The ascending aorta was larger than the descending aorta, but showed similar stiffness range. However, it should be noted that the distensibility values measured and reported here are the apparent in-vivo ones. These are measured in fixed 2D planes, without accounting for the longitudinal deformation. Indeed, in ascending aorta, the left ventricle contraction causes a longitudinal stretch which may lead to a small cross-sectional diameter and thereby cause the observed stiffness to be higher [47–49]. In contrast, the descending aorta’s length remains largely fixed over cardiac cycle.

Even though, overall, there was a clear difference between the aortic diameter in males and females, it is largely explained by the height/weight between males and females. That is, after accounting for the BSA, the aortic size difference between males and females becomes much smaller, although not zero. Moreover, there was no sex-based difference in the aortic stiffness. These findings may have implications for the clinical guidelines on aortic disease management. These findings can be used to inform aortic device design where compliance matching is desired, for example when developing different devices for varying BSAs or males/females.

Patient-specific computational models and digital twins are highly promising for improving the diagnosis and treatment [50–52]. However, in the current state-of-the-art, only the geometry is the main personalised feature, while the stiffness parameters used tend to be merely generic values. Based on the above findings, we can now choose aortic stiffness more representative of a specific patient, based on their age, sex, height and weight, blood pressure, and even aortic diameter. This could be done by using only the mean values or also the observed variation to establish confidence in the results, thus significantly improving the patient-specific nature of our models and their credibility.

### 4.2 Main findings on the stresses

One of the primary motivations for estimating stresses in the aortic tissue is to assess the risk of adverse events, such as aortic dissection and rupture. In contrast to stiffness, which can be measured in-vivo, stresses can only be computed based on a model. A common way to estimate the circumferential stress is based on the law of Laplace [53, 54]. According to which, circumferential stresses are assumed to be uniform across the wall and given by *PD/*2*h*, where *P* is the pressure, *D* is the diameter, and *h* is the tissue thickness. For systole and diastole, one would use the corresponding values of pressure, diameter and thickness. However, this traditional approach has three main limitations: 1) it does not account for the relation between pressure, diameter, and stiffness; 2) it assumes uniform stress across the wall, and ignores the effect of non-uniform distribution; and 3) it does not provide any information on the longitudinal stresses.

Herein, an innovative approach was used, which integrated the extensive ex-vivo information on arterial tissues with in-vivo observations to address the limitations of the law of Laplace approach. The thick-walled model allowed us to compute detailed distributions of stresses across the arterial wall, and in all three directions. In addition to the expected values, the associated uncertainty was also quantified.

The stresses in circumferential and longitudinal direction were mostly tensile, and of the same order, while the radial stress was compressive and of much smaller value (Figs. 7,8). The stress in the circumferential direction at the inner wall was generally the highest and significantly amplified as compared to the lumen pressure (sometimes as high as 100 times the lumen pressure). In fact, it has been previously proposed that the presence of residual stresses in the aortic tissue helps to reduce such stress concentrations [55]. Here, even though the residual stresses (in terms of opening angle) were incorporated, high stress values were still observed. Incorporating this hypothesis into the proposed inverse framework, one could optimise the opening angle such that the stress concentrations are minimised. Even after normalising the stresses with respect to the lumen pressure, the circumferential stress at the inner wall at systole was found to be higher than that at diastole. This is primarily due to the nonlinear stiffening behaviour of the aortic tissue.

Importantly, if the high stress values exceed the tissue strength (i.e., the maximum stress that the aortic tissue can carry), it will likely result in the initiation of an aortic dissection. As a rough comparison, the tissue strength for diseased ascending aortic tissue has been reported as 1.510 ± 1.030 MPa and 0.587 ± 0.232 MPa in the circumferential and longitudinal directions, respectively [56]. When normalised with respect to a mean SBP of 120 mmHg, we obtain a range of 94.4 ± 64.4 and 36.7 ± 14.5 for the normalised tissue strength. Thus, the calculated stresses are just below the diseased tissue strength range. In fact, it is straightforward to compare the two (calculated stress and strength) to calculate a patient-specific “dissection risk”, which can inform clinical decision-making. However, since the strength is also expected to vary depending on patient-specific factors (such as disease), this extension will be pursued with more data on aortic strength in the future.

Varying the in-vivo observations, it was found that the stress values increased with the aortic diameter (Fig. 9c). This result is consistent with the law of Laplace. A noteworthy finding is the observed decrease in stress with distensibility (Fig. 9d), which is novel and significant. This was observed for both diastole and systole, and in both circumferential and longitudinal directions. Recall that with age distensibility decreases and aortic size increases (Fig. 6a). Thus, combined together, the two results imply that the maximum stress (and thereby the risk of adverse events) may compound and increase super-linearly with age.

It is also worthwhile comparing the observed trends in stresses with respect to distensibility against the predictions based on law of Laplace. The law of Laplace implies that, as distensibility increases, there is no change in diastolic stress, while the systolic stress should increase (since higher distensibility means larger systolic diameter and lower systolic thickness). However that does not take into account the negative correlation between diameter and distensibility. Indeed, the results here showed a negative correlation between the distensibility and systolic *mean stress* (which is equivalent to the stress calculated using law of Laplace (Fig. 10). Interestingly the correlation between distensibility and *maximum* stress becomes even more strongly negative, even after accounting for the associated uncertainty. These results emphasise the advantage of the proposed approach, as compared to the law of Laplace.

Current clinical guidelines use only the aortic diameter to assess the risk of adverse events, which is partly guided by the law of Laplace. However, the results presented here indicate that distensibility also plays an important role in the stresses. It was further confirmed using MI (Fig. 11), which showed a significant increase when both diameter and distensibility were used together. In fact, the improvement from incorporating distensibility was almost the same as that resulting from inclusion of tissue thickness. Previous studies have supported the inclusion of distensibility in the risk assessment [56, 57], and the results presented here further strengthen that argument.

Another limitation of the current clinical guidelines is that the tissue thickness is not accounted for. It is natural to expect that a thicker arterial wall will experience lower stresses, which is also supported by the law of Laplace. The results presented here were consistent, with negative correlation coefficients – in both circumferential and longitudinal directions (Fig. 10). Similarly, the mutual information results here suggest that inclusion of tissue thickness can improve the clinical guidelines. Although observing tissue thickness in-vivo is challenging (it was not possible from the CMR used here), the upcoming advanced imaging techniques, such as photon counting CT [58], may help overcome that limitation.

### 4.3 Limitations and future directions

The presented approach carries some limitations related to the imaging and segmentation methods. The location and angle of the CMR plane may affect the aortic size calculations. The automatic segmentation method may also introduce errors, both due to the limited image resolution and the inaccurate prediction in some cases. The latter were largely removed through quality control testing. Similarly the conversion from lumen area to diameter assumes circular shape, which may not always be strictly true. The UK Biobank cohort is also expected to be biased towards certain health and socioeconomic factors [59], and the proposed framework should be tested against another large dataset. Some of these limitations will be improved upon in the future.

The aortic properties may also depend on other demographic factors, such as exercise and smoking habits. These were not included in the current study, but the proposed framework is easily extendible to study any such effects, and will be investigated in the future. More broadly, the proposed framework of using statistical models, such as GMMs, to model joint distributions and analysing the conditional distributions can be applied to other features and datasets.

An important limitation of the stress calculations was that there was no information on the longitudinal deformation (due to 2D nature of the CMR). This contributed to the high uncertainty in longitudinal stresses, and innovative approaches are required to address this limitation. It could be partially addressed by using another, longitudinal CMR view. However, that view will still not provide information on the ex-vivo to in-vivo stretch, at least directly, and further work is required to reduce the uncertainty in the predicted longitudinal stresses.

The model employed could be further refined, e.g. in terms of layer specificity. Overall, the proposed framework to model the relationship between the in-vivo observations and the resulting wall stresses can be viewed as a digital twin, which allows fast prediction of arterial biomechanical state. Adding more realism into these models will pave the way towards biomechanically-informed clinical decision making.

### 4.4 Conclusion

This study presents the *largest population-scale* characterisation of human aortic biomechanics ever performed, and the results provide crucial information about human aortic biomechanical properties and their trends and variations. The insights gained will help improve the personalisation of computational models, drive the clinical guidelines, and provide a baseline to compare diseased patients against. Thus, this is a new reference framework for personalised biomechanical assessment of the thoracic aorta, which, in the future, will be further extended to investigate other aspects of aortic biomechanics, including its longitudinal deformation and effect of three-dimensional shape.

## Supporting information

Animation of distensibility with age

Animation of distensibility with DBP

Animation of distensibility with height

Animation of distensibility with SBP

Animation of stress with DBP

Animation of stress with diameter

Animation of stress with distensibility

Animation of stress with SBP

Animation of distensibility with weight

## Data Availability

All data produced in the present study are available upon reasonable request to the authors

## Supplementary information

Animated plots of conditional distributions of size, stiffness, and stresses are provided as supplementary information.

## Acknowledgements

This work was partially supported by the Engineering and Physical Sciences Research Council of the UK (grant number EP/X5257161/1). We also thank the support from the University of Glasgow’s Global Partnership Fund. This research has been conducted using the UK Biobank Resource under application number 169530.

## Author contributions

Conceptualisation: A.A., C.C., S.S. Methodology: A.A., J-E.B. Software: A.A. Visualisation: A.A. Formal analysis: A.A. Investigation: A.A., C.C., M.D., N.F., A.P. Funding acquisition: A.A., A.McB., M.D. Writing–original draft: A.A. Writing–review and editing: all authors.

## Conflict of interest

Authors declare there is no conflict of interest.

## Appendix A Mutual information

Mutual information (MI) quantifies the statistical dependence between two random variables by measuring how much knowledge of one variable reduces uncertainty about the other. It is defined in terms of the joint distribution *p*_(*X,Y*)_ and the associated marginal distributions *p*_*X*_ (*x*) and *p*_*Y*_ (*y*) as

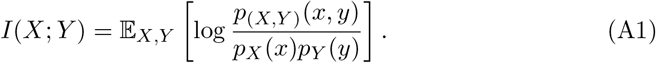

Mutual information is non-negative and equals zero if and only if *X* and *Y* are statistically independent. Unlike linear correlation measures, MI captures arbitrary nonlinear dependencies.

The MI was calculated following an approach by [34], which minimises the bias through a non-parametric *k*-nearest neighbour entropy estimator. Python library npeet was used for the MI computation.

### A.1 Conditional mutual information

A useful extension of MI is the conditional mutual information (CMI), which defines the mutual information between two variables *X* and *Y*, while a third variable *Z* is given/known. CMI, denoted as *I*(*X*; *Y*| *Z*), can be used to find second-order effects, where the MI between *X* and *Y* might be only because of *Z*.

While <monospace>npeet</monospace> supports mixed-variable MI, it does not provide a general estimator for CMI between a continuous and a discrete variable (e.g., if *Y* is discrete and (*X, Z*) are continuous). Therefore, an alternative model-based approach was used for estimating CMI in the mixed-variable setting. The details of this approach are deferred until after the models employed have been defined.

## Appendix B Gaussian mixture model fitting

Gaussian mixture models (GMMs) provide a flexible parametric framework for modelling multivariate continuous probability distributions. A GMM represents a density as a convex combination of multivariate Gaussian components, allowing complex, multimodal distributions to be approximated.

Let *X*∈ R^*d*^ denote a continuous random vector. A Gaussian mixture model with *K* components defines the density

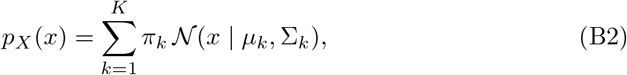

where *π*_*k*_ ≥ 0 are mixture weights satisfying Σ_*k*_ *π*_*k*_ = 1, and N(*x* | *µ*_*k*_, Σ_*k*_) denotes a multivariate normal distribution with mean vector *µ*_*k*_ and covariance matrix Σ_*k*_.

The GMM model was fitted to a multi-dimensional dataset of continuous variables, with a train/test split of 75/25. Since most of the variables in this study are strictly positive, a log-transform was tested, if appropriate. Model parameters 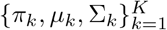 were estimated using the expectation–maximisation (EM) algorithm on the training subset. The number of mixture components *K* was selected to minimise the Bayesian Information Criterion (BIC) on the testing subset. Similarly, the appropriateness of the log-transform was confirmed by comparing the BIC values.

### B.1 Joint modelling of multiple variables

When modelling multiple continuous variables jointly, for example

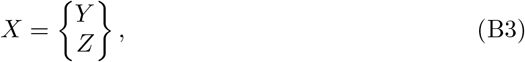

a GMM is fitted to the combined variable *X*, yielding a density of the form

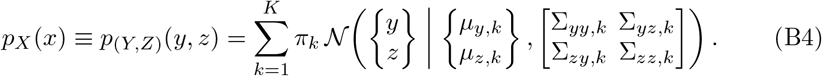

This joint modelling allows dependencies between variables to be captured within each mixture component and across the mixture as a whole.

#### B.1.1 Marginal distributions from a GMM

Marginal distributions of a subset of variables can be obtained analytically from a fitted GMM. If the joint density *p*_(*Y,Z*)_(*y, z*) is modelled by a GMM, the marginal density of *Z* is given by

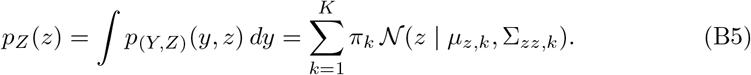

Thus, the marginal distribution remains a Gaussian mixture with the same mixture weights, where each component’s mean and covariance correspond to the relevant sub-blocks of the original joint mean vector and covariance matrix, respectively.

##### B.1.2 Conditional distributions from a GMM

Conditional distributions can also be computed in closed form. For a single Gaussian component, the conditional distribution of *Y* given *Z* = *z* is Gaussian:

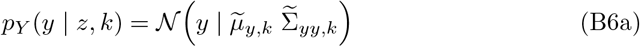

Where

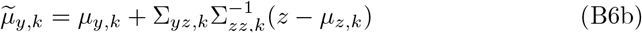

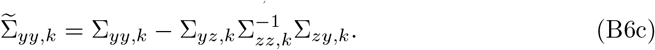

For the full mixture model, the conditional distribution *p*_*Y*_ (*y* | *z*) is itself a Gaussian mixture:

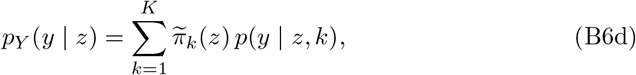

where the conditional mixture weights depend on the observed value *z*:

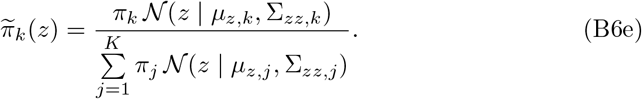

All marginal and conditional quantities required for downstream analysis can therefore be computed analytically once the joint GMM has been fitted. This property allows GMMs to be used as flexible probability models, providing distributions over one set of variables conditioned on observed values of another.

#### B.2 GMM-based CMI estimation for mixed variables

To compute CMI between a continuous variable *X* and a discrete variable *Y* given a continuous variable *Z*, GMM-based estimation was used. Separate GMMs were fitted to the joint continuous variables (*X, Z*) for each value of the discrete variable *Y*, yielding class-conditional GMM for densities *p*(*x, z*|*y*). Class prior probabilities *p*(*y*) were estimated empirically. The approach relies on the identity

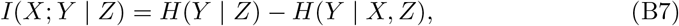

where *H*(·) denotes Shannon entropy, which in-turn relies on the conditional probabilities of the discrete variable *Y* : *p*(*y* | *z*) and *p*(*y*|*x, z*).

These conditional probabilities were computed using the fitted GMM and Bayes’ rule. That is, for a given observation (*x, z*),

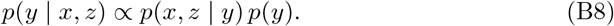

Similarly, the posterior distribution of *Y* given *Z* alone is obtained by first marginalising the class-conditional GMMs over *X* to get *p*(*z* | *y*), followed by the Bayes’ rule:

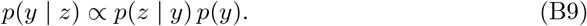

The conditional entropies are estimated by Monte Carlo integration over the empirical data distribution,

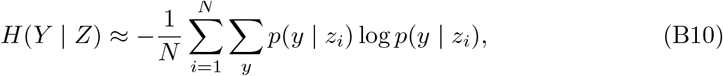

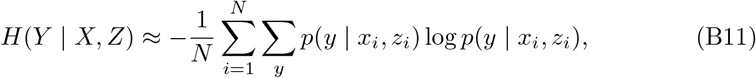

where the expectation was approximated using the combined dataset from all classes. Finally, the CMI was calculated using the identity (B7).

## Appendix C Biomechanical modelling of the aorta

Aorta was modelled as a thick elastic cylinder that deforms under an internal pressure [26, 60]. If the reference configuration is denoted as Ω_R_, the aim is to find the deformed configuration denoted as Ω_D_ such that it is in equilibrium with the applied loads and boundary conditions. This problem can be framed in terms of the mapping between the two configurations. Importantly, the presence of residual stresses means that the “true” reference configuration is significantly different from in-vivo state, thus necessitating fully nonlinear kinematic description. The residual stresses have been demonstrated through opening angle (O.A., *ω*) and longitudinal stretch *λ*_*Z*_.

If we assume axisymmetric deformation of a perfect cylinder, this problem reduces to finding a map between the radii (undeformed radius *R* and deformed radius *r*). Moreover, incompressibility implies that the deformed radius at any point can be expressed in terms of one (scalar) deformation measure, such as the internal radius— undeformed *R*_*i*_ and deformed *r*_*i*_ [60]. That is, the mapping can be written as:

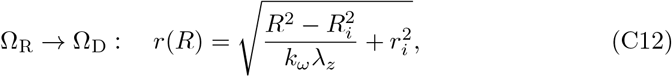

where *k*_*ω*_ = 2*π/*(2*π* − *ω*), and the deformation gradient becomes

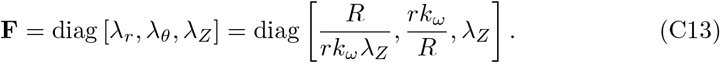

The pressure-deformation still follows the same relationship (C16) but with modified stretches (C13). It should be noted that *R*_*i*_ and *H* here represent the *ex-vivo* radius and thickness, not *in-vivo*, as the two can be significantly different.

Given a deformation gradient, **F**, the stresses are derived from a constitutive model. For arterial tissue, a hyperelastic model proposed by Gasser–Ogden–Holzapfel (GOH) [61] is widely used and adopted here. It defines the strain energy density Ψ as:

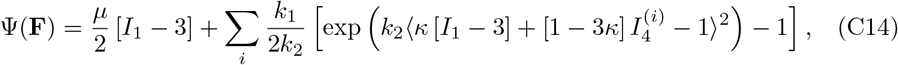

where *I*_1_ = tr **F**^⊤^**F** is the first strain invariant and 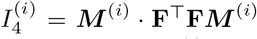 is the pseudo-invariant representing stretch squared along (fibre) direction ***M*** ^(*i*)^. The tissue was assumed to be made up of two fibre families symmetrically arranged in a helical fashion, about the circumferential direction at angles ± *ϕ*, each with a direction vector ***M*** ^(*i*)^. *κ* is a dimensionless parameter representing the fibre dispersion, and ⟨·⟩denotes the Macaulay bracket.

From the strain energy density, the Cauchy stress tensor is derived as:

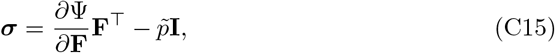

where 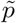 is the Lagrange multiplier used to enforce incompressibility (det(**F**) = 1). For a diagonal deformation gradient (Eq. C13) and symmetric fibre directions (at ±*ϕ*), the Cauchy stress tensor also comes out to be diagonal, with three normal stresses: radial stress *σ*_*rr*_, circumferential stress *σ*_*θθ*_, and longitudinal stress *σ*_*zz*_, while the shear stresses are zero.

Lastly, the stresses must satisfy equilibrium, which entails the following balance between the lumen pressure *P* and internal stresses:

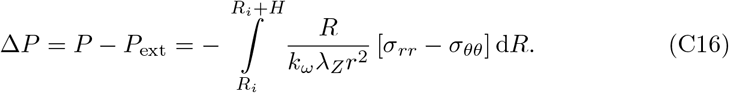

Here *H* is the arterial wall thickness in the stress-free configuration. The external pressure *P*_ext_ is assumed to be zero. Thus, the lumen pressure required to equilibrate a deformed internal radius *r*_*i*_ can be calculated. Inversely, the deformed radius at a given lumen pressure can also be determined numerically. Finally, at a given lumen pressure (or correspondingly, a given deformed radius), the Lagrange multiplier 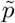 and Cauchy stress variation across the arterial wall are calculated. These calculations were performed using an open-source Python library, pyMechT [62].

## Appendix D Stress results for descending aorta

Calculated stresses for the descending aorta are shown in Figures D1, D2, and D3. Qualitatively, these show the same behaviour as the ascending aorta.

**Fig. D1.**
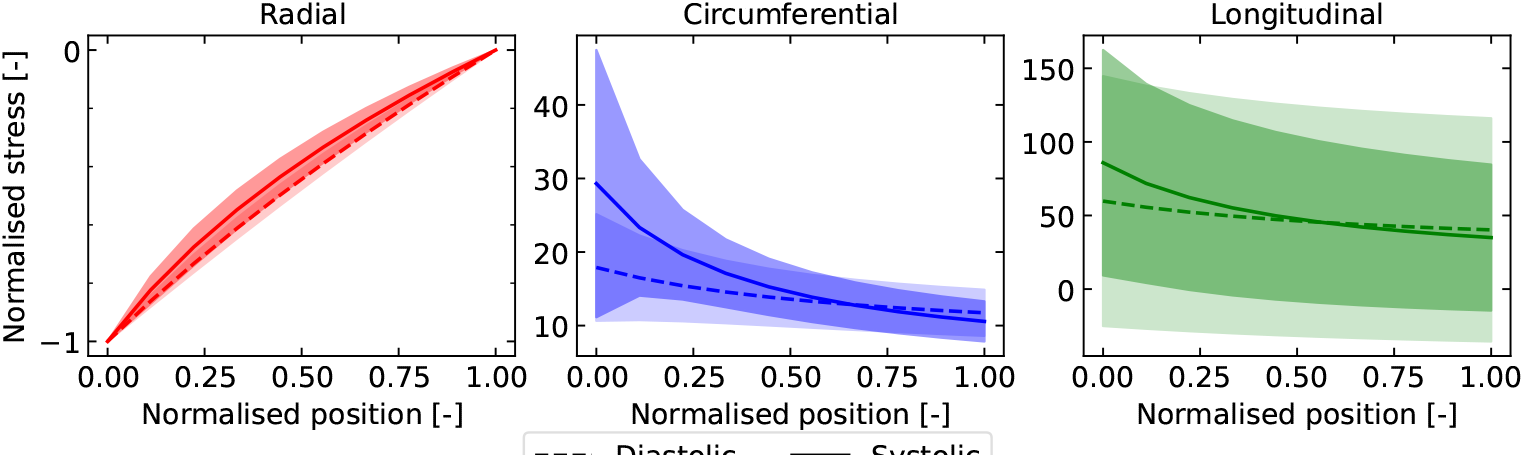
Distribution of the normal, circumferential, and longitudinal stresses (normalised to the blood pressure) in the descending aorta across the arterial wall (normalised position, 0 being the inner surface and 1 being the outer), with variation for all cases combined (i.e., marginalised to the aortic parameters). Solid lines represent the mean values and shaded areas indicate one standard deviation. Radial stresses are compressive with small uncertainty, while the circumferential and longitudinal stresses are predominantly tensile with high uncertainties.

**Fig. D2.**
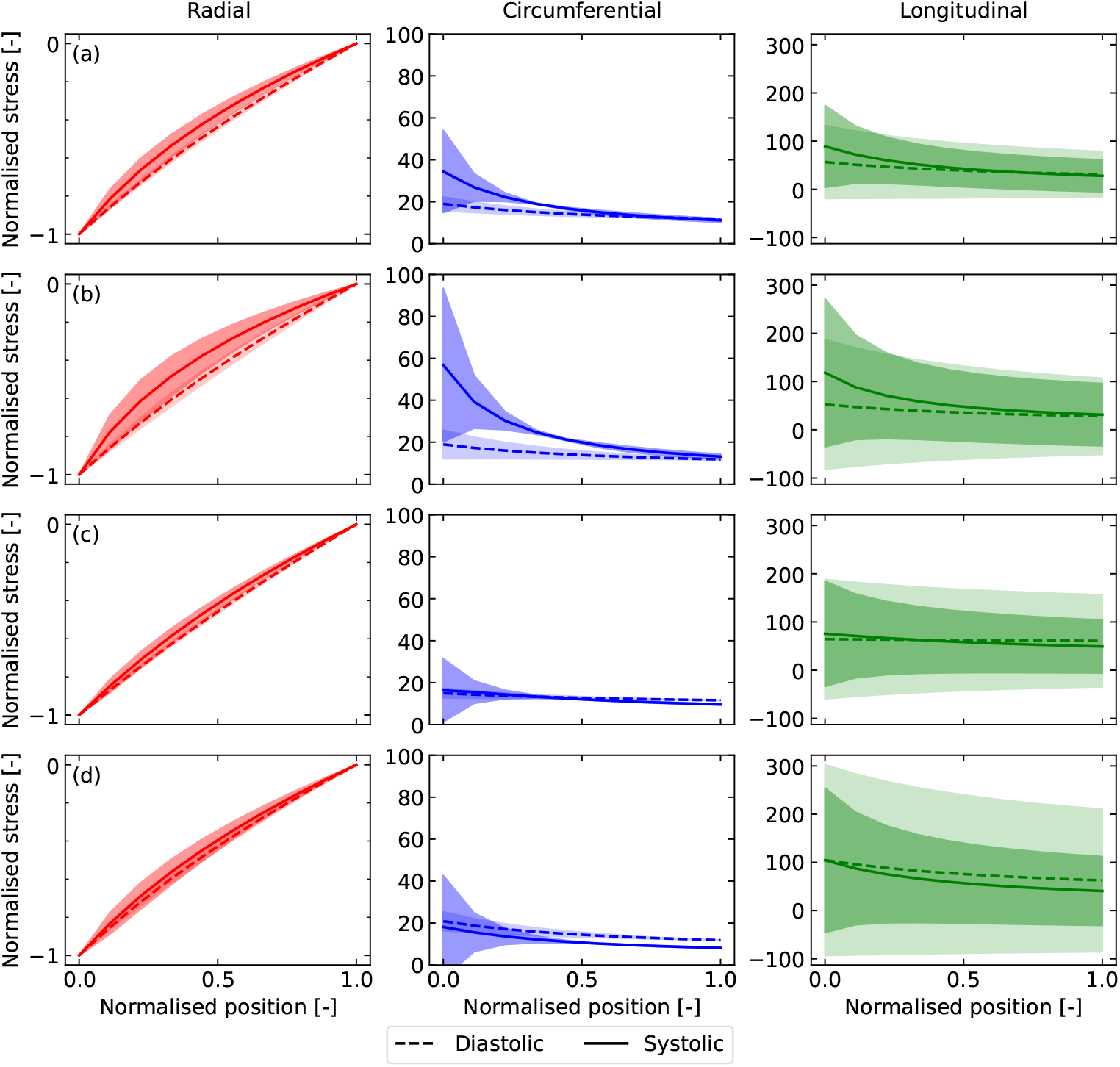
Example conditional distributions of the stresses (normalised to the blood pressure) across the descending arterial wall (normalised position, 0 being the inner surface and 1 being the outer) for different blood pressures, aortic size and distensibility: a) DBP = 65 mmHg, SBP = 130 mmHg, diameter = 25 mm, dist = 2 × 10^−3^ mmHg^−1^; b) DBP = 80 mmHg, SBP = 130 mmHg, diameter = 30 mm, dist = 3 × 10^−3^ mmHg^−1^; c) DBP = 55 mmHg, SBP = 120 mmHg, diameter = 20 mm, dist = 4 × 10^−3^ mmHg^−1^; d) DBP = 75 mmHg, SBP = 150 mmHg, diameter = 20 mm, dist = 1 × 10^−3^ mmHg^−1^. Solid/dashed lines are the mean values and shaded areas represent one standard deviation.

**Fig. D3.**
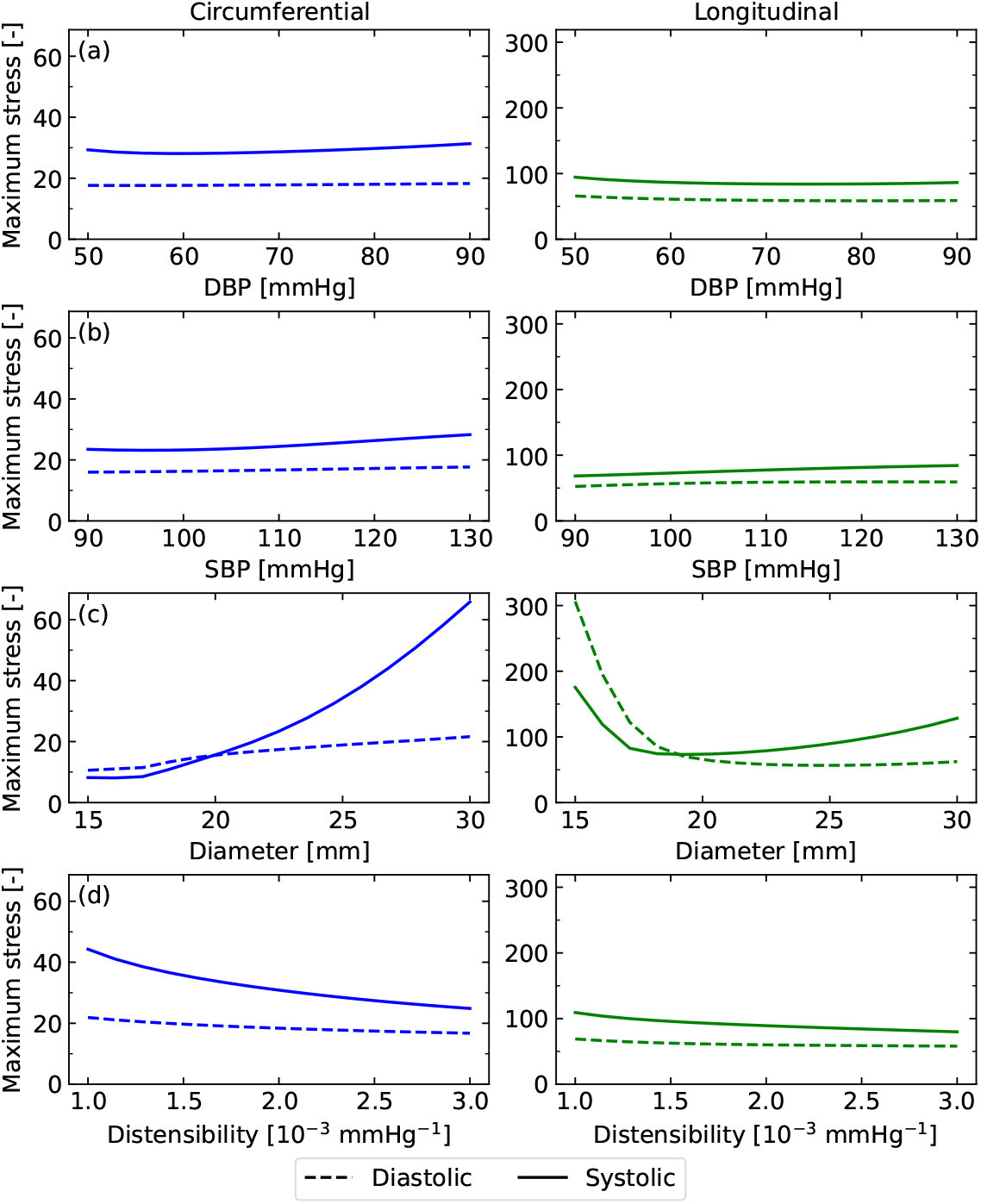
Variation of the expected maximum stress (normalised) as the pressures, size and distensibility of the descending aorta are varied. Pressures have a minimal effect, however the stress increases with diameter, but decreases with distensibility.

## Notes

### Competing Interest Statement

The authors have declared no competing interest.

### Author Declarations

The study used ONLY openly available human data that were originally located at UK Biobank

