## Supplementary figures and images for "Population-scale trends and variations in human aortic biomechanics"

### Animation of distensibility with age

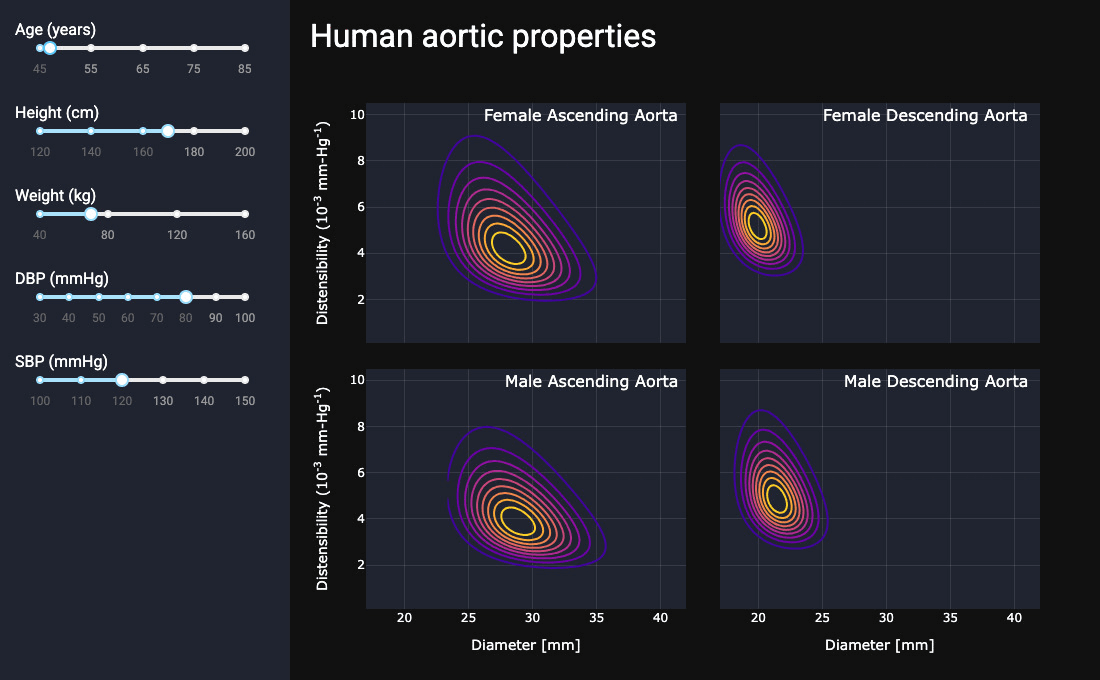

### Animation of distensibility with DBP

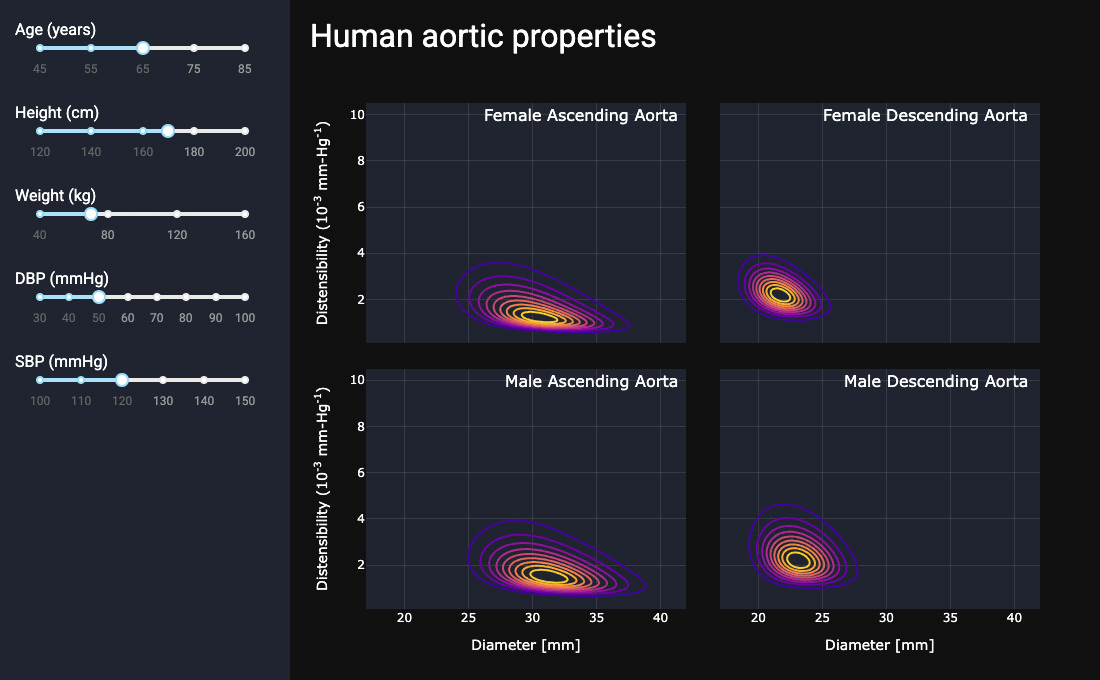

### Animation of distensibility with height

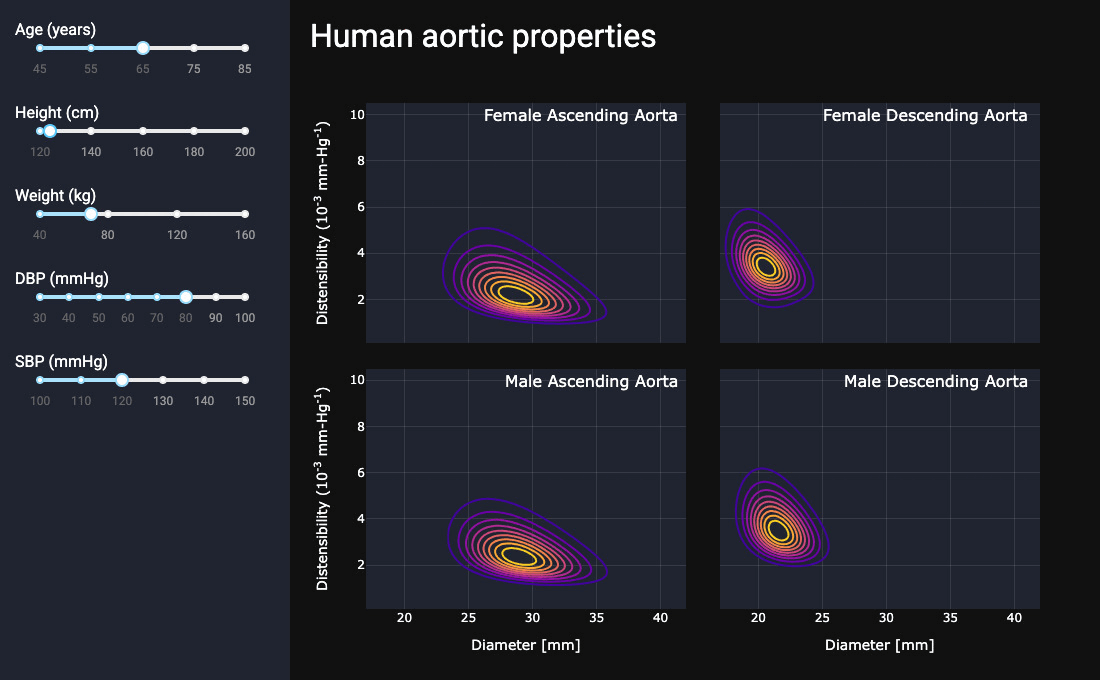

### Animation of distensibility with SBP

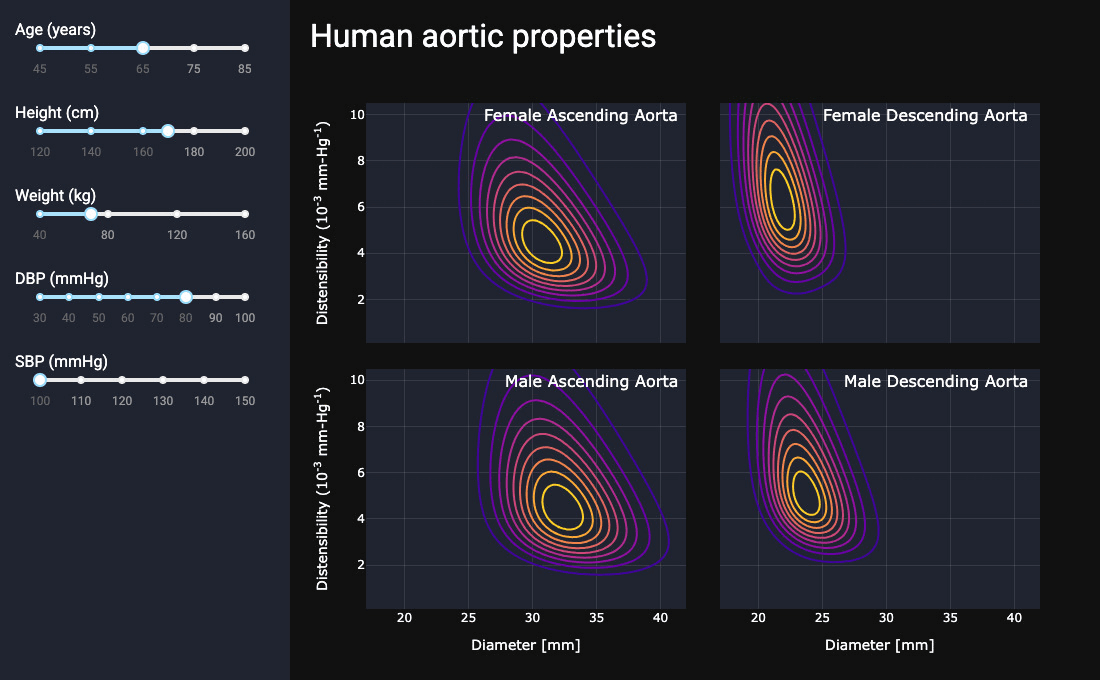

### Animation of distensibility with weight

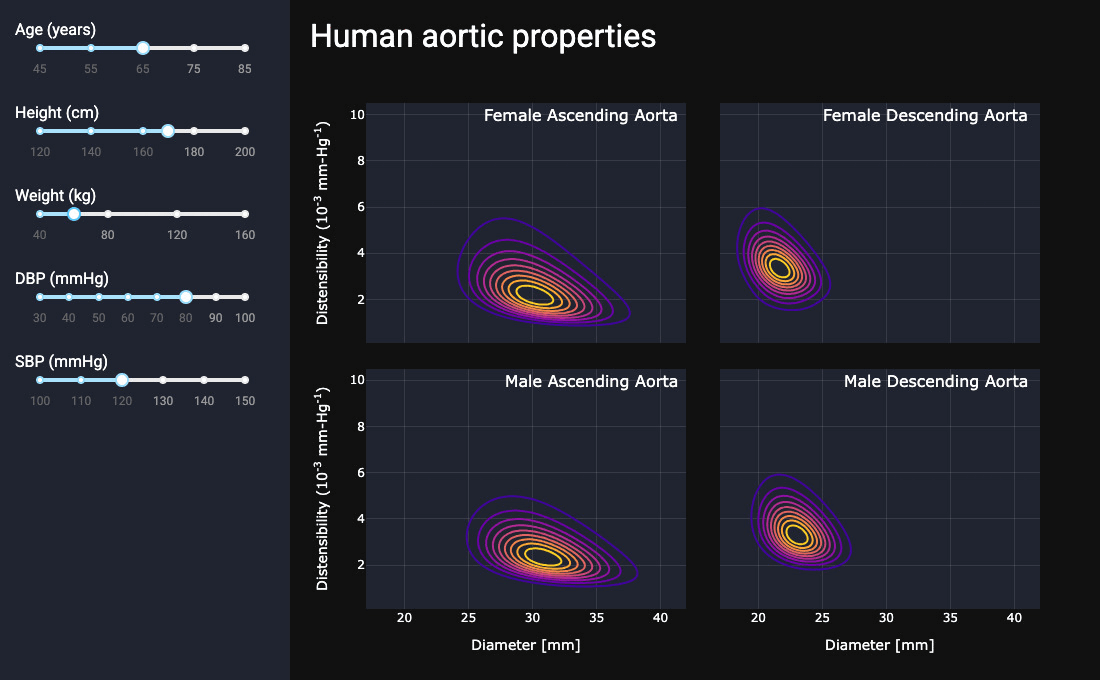

### Animation of stress with DBP

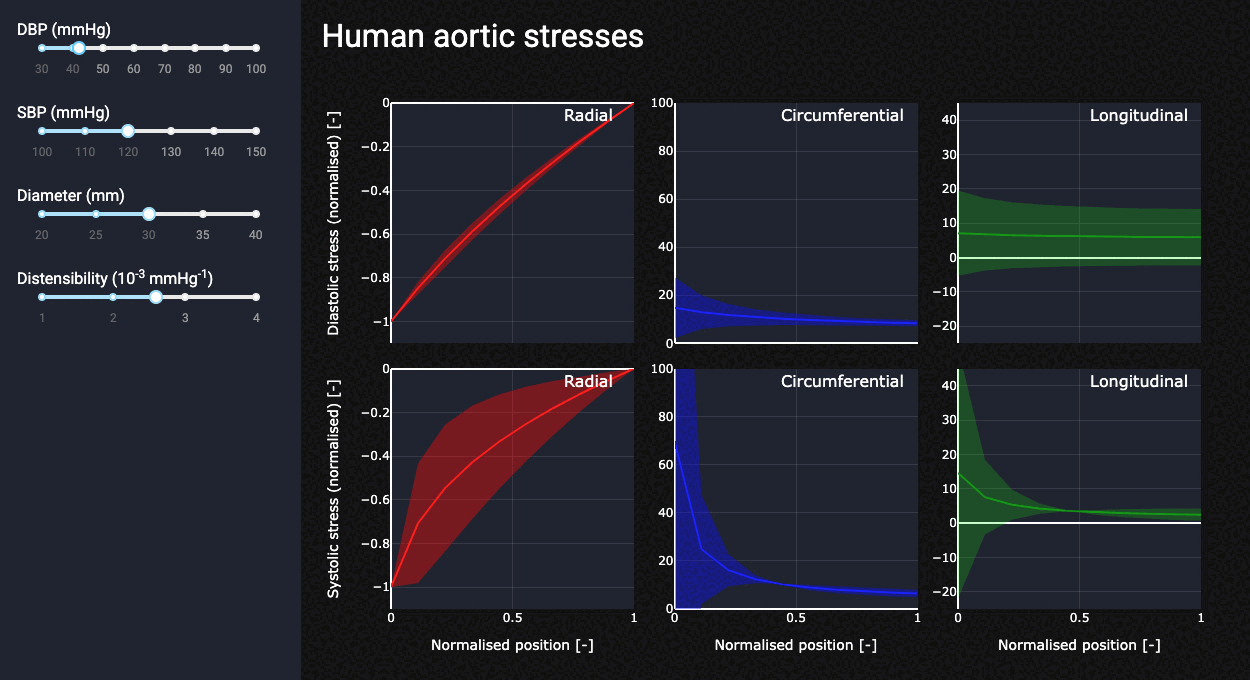

### Animation of stress with diameter

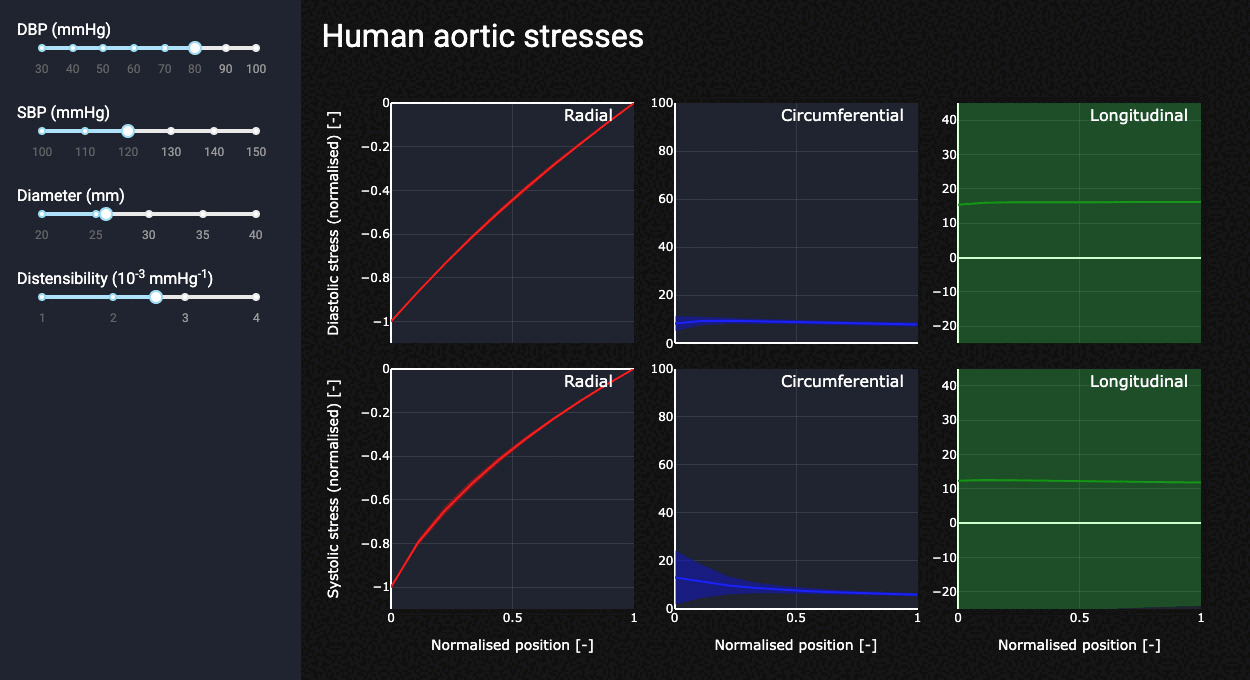

### Animation of stress with distensibility

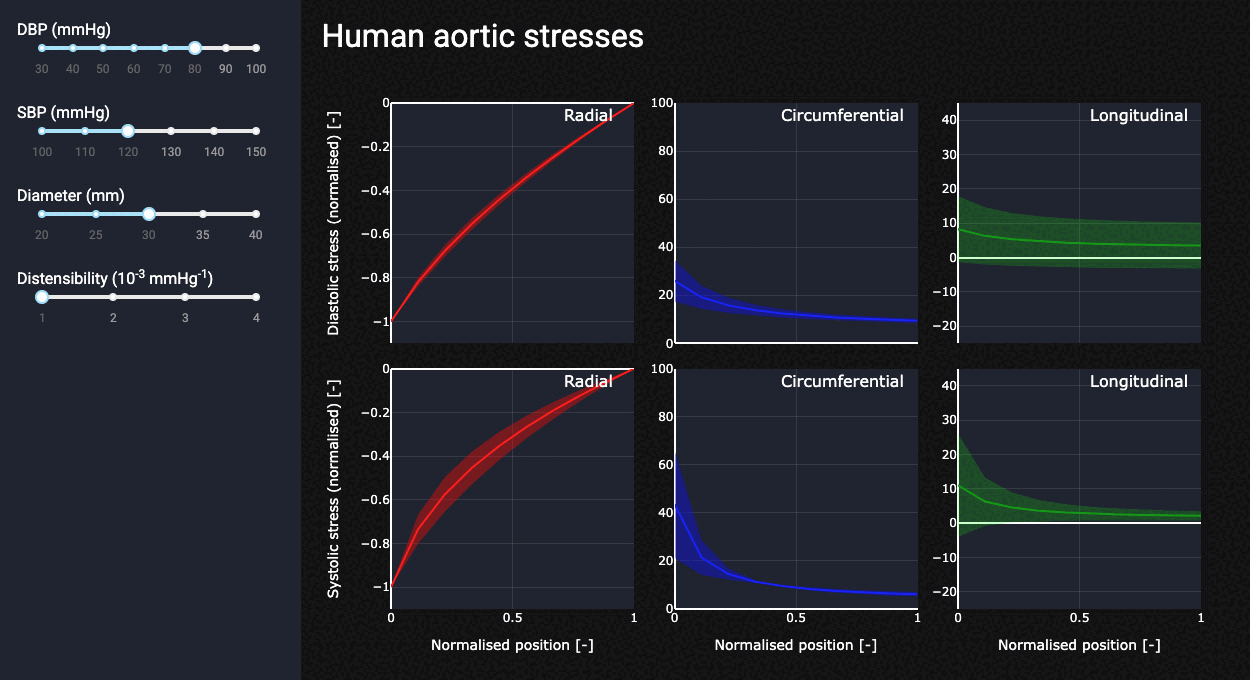

### Animation of stress with SBP

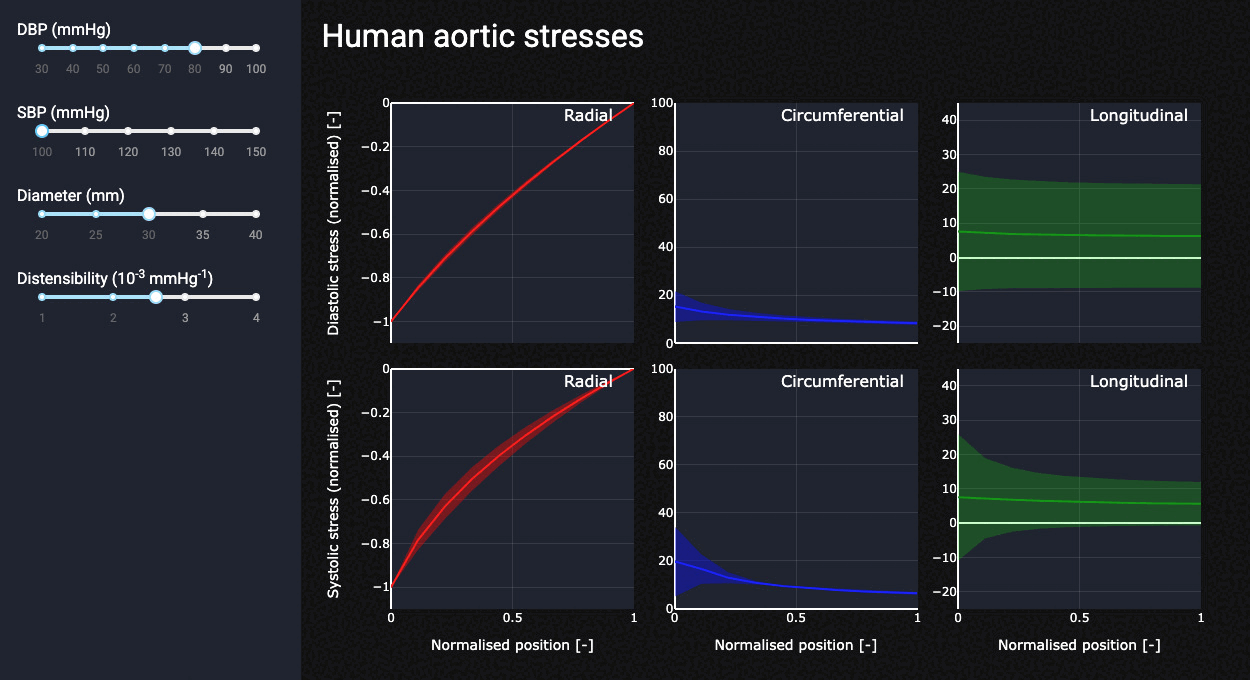
